# Identifying leptospirosis hotspots in Fiji using a One Health model that incorporates watershed-scale pathogen transport

**DOI:** 10.64898/2026.08.03.26359543

**Authors:** Ama R. Wakwella, Carissa J. Klein, Owen Woodberry, Colleen L. Lau, Amelia Wenger, Stacy D. Jupiter, Aaron P. Jenkins, Helen J. Mayfield

**Affiliations:** University of Queensland, School of the Environment, Faculty of Science, St Lucia QLD 4072, Australia; Centre for Biodiversity and Conservation Science, The University of Queensland, St Lucia, QLD, 4072, Australia; Monash University, Faculty of Information Technology, Monash Data Futures Institute, VIC 3800, Australia; University of Queensland, Frazer Institute, Faculty of Health, Medicine, and Behavioural Sciences, Herston, QLD 4029, Australia; Wildlife Conservation Society, Global Marine Program, Bronx, NY 10460, USA; The University of Sydney, Sydney School of Public Health, Sydney Institute for Infectious Diseases, Camperdown NSW 2006, Australia; Edith Cowan University, School of Science, Centre for People Place and Planet, Joondalup WA 6027, Australia

**Keywords:** Infectious disease modelling, catchment, One Health, epidemiology, land use, Pacific Islands

## Abstract

Leptospirosis is a water-related zoonotic disease with complex transmission pathways, including direct transmission from infected animals and indirect transmission through contaminated soil and water. Identifying key areas to implement targeted infection prevention and control strategies is challenging, as a range of risk factors across different scales can drive human infection. We aimed to develop an epidemiological modelling approach to predict key transmission pathways driving leptospirosis infection, including risks ranging from household level factors to the movement of pathogens across watersheds. We combined a causal Bayesian network with a novel hydrological pathogen transport model to predict leptospirosis across Fiji and found key infection hotspots adjacent to rivers and within degraded watersheds; a dynamic overlooked by previous epidemiological models. We used a wide range of data for model parameterisation (e.g., expert elicitation, epidemiological surveys, and environmental data) and found that predictive validity improved when expert input on risk factors was used to guide model parameterisation, improving R^2^ for predicted versus observed seroprevalence from 0.71 to 0.91. Our One Health modelling approach can be used to support the design and evaluation of environment-based disease prevention strategies at national scales.

## Introduction

Leptospirosis is a zoonotic, water-related infectious disease responsible for severe health burdens across the globe. This includes an estimated 58,900 deaths (1), 2.9 million lost disability-adjusted life years (2), and the loss of billions of dollars due to loss of productivity in humans every year (3). Despite the potential global benefits, the development of targeted leptospirosis prevention and control strategies remains hindered by both limited disease surveillance systems (4–6), as well as gaps in our understanding of the complex transmission pathways that drive infections (7, 8).

Leptospirosis transmission pathways include complex interactions between people, animals, and the environment due to the broad infection and survival capabilities of the pathogenic *Leptospira* bacteria (9). Although humans rarely transmit leptospirosis, many other animals (e.g., domestic animals, livestock, and wildlife) facilitate transmission by excreting pathogenic *Leptospira* through their urine and into the environment, where the bacteria can remain viable for months (9, 10). Infection in humans occurs either directly through contact with infected animals, or indirectly through exposure to contaminated soil or water. Rainfall-induced transport of the bacteria throughout a watershed, via the erosion and runoff of contaminated soils, further complicates effective interventions (8, 10, 11) as pathogens could originate from animals either within communities or upstream. While a range of socio-demographic and environmental risk factors also affect the likelihood of human exposure to *Leptospira* (9), the impact of these drivers can vary greatly between regions, and their relative importance on disease burden is often unclear (12, 13).

The Pacific Islands have some of the highest health burdens from leptospirosis in the world (1). In Fiji where leptospirosis has been listed as a top priority for management within Fiji’s Ministry of Health’s Strategic Plan for 2020-2025 (14), epidemiological models have been developed to assist in predicting leptospirosis and/or evaluating risk factors for infection (15–21). However, as in other countries, these models make limited distinction between direct and indirect transmission (22, 23) and to date, there has been no explicit evaluation of how pathogenic *Leptospira* enters and is transported across watersheds through surface runoff (8, 10, 24).

The use of a One Health approach that acknowledges the intrinsic links between animals, environments, and humans is increasingly recommended as a framework for epidemiological modelling of zoonotic diseases, such as leptospirosis (25, 26). Applying a One Health framework in Fiji to integrate socio-demographic, animal host, and environmental drivers – including pathogen transport across watersheds – could help address key gaps in understanding regional transmission pathways. Crucially, this approach also enables management actions to be planned and evaluated across multiple scales (e.g., household and upstream (27, 28)).

Here, we aim to model the direct and indirect transmission pathways driving leptospirosis infection in Fiji using a hybrid approach that allows for movement of pathogens across watersheds to be considered. We build upon previous leptospirosis models by combining a causal Bayesian network (BN) with the results of a newly developed hydrological pathogen transport model. This novel approach allows the model to also consider the risk from *Leptospira* entering the environment upstream, as well as local factors that contribute to exposure.

## Methods

### Study region

Our study region is four major islands of the Fiji archipelago (Viti Levu, Vanua Levu, Taveuni, and Ovalau), a tropical region in the southern Pacific Ocean with a total population of approximately 906,124 people (29). The majority of the population is *iTaukei* (Indigenous Fijian; ∼57 %) or Fijian of Indian descent (Indo-Fijians; ∼38 %; (30)). Although over half of the country’s population engages in subsistence farming (31), contact with different livestock species can vary significantly between ethnic groups and different residential settings (16). The most recent cross-sectional survey of leptospirosis infection in Fiji in 2013 found that approximately 19.4 % of people were seropositive (had *Leptospira* antibodies in their blood), indicating past or present leptospirosis infection (15).

### Modelling overview

We used an ensemble modelling approach combining a spatially explicit BN with a novel hydrological pathogen transport model to explore risk factors that influence leptospirosis seropositivity at the community level. The BN structure was designed using an iterative approach to integrate expert knowledge, household survey data, and evidence from published literature, creating a causal representation of direct and indirect transmission pathways in Fiji (Fig 1). The probabilities for the BN were quantified by integrating the outputs of the hydrological pathogen transport model, together with published sources and expert knowledge. Model predictions were used to generate maps of predicted leptospirosis seroprevalence and the likelihood of direct or indirect transmission across Fiji.

**Fig 1.**
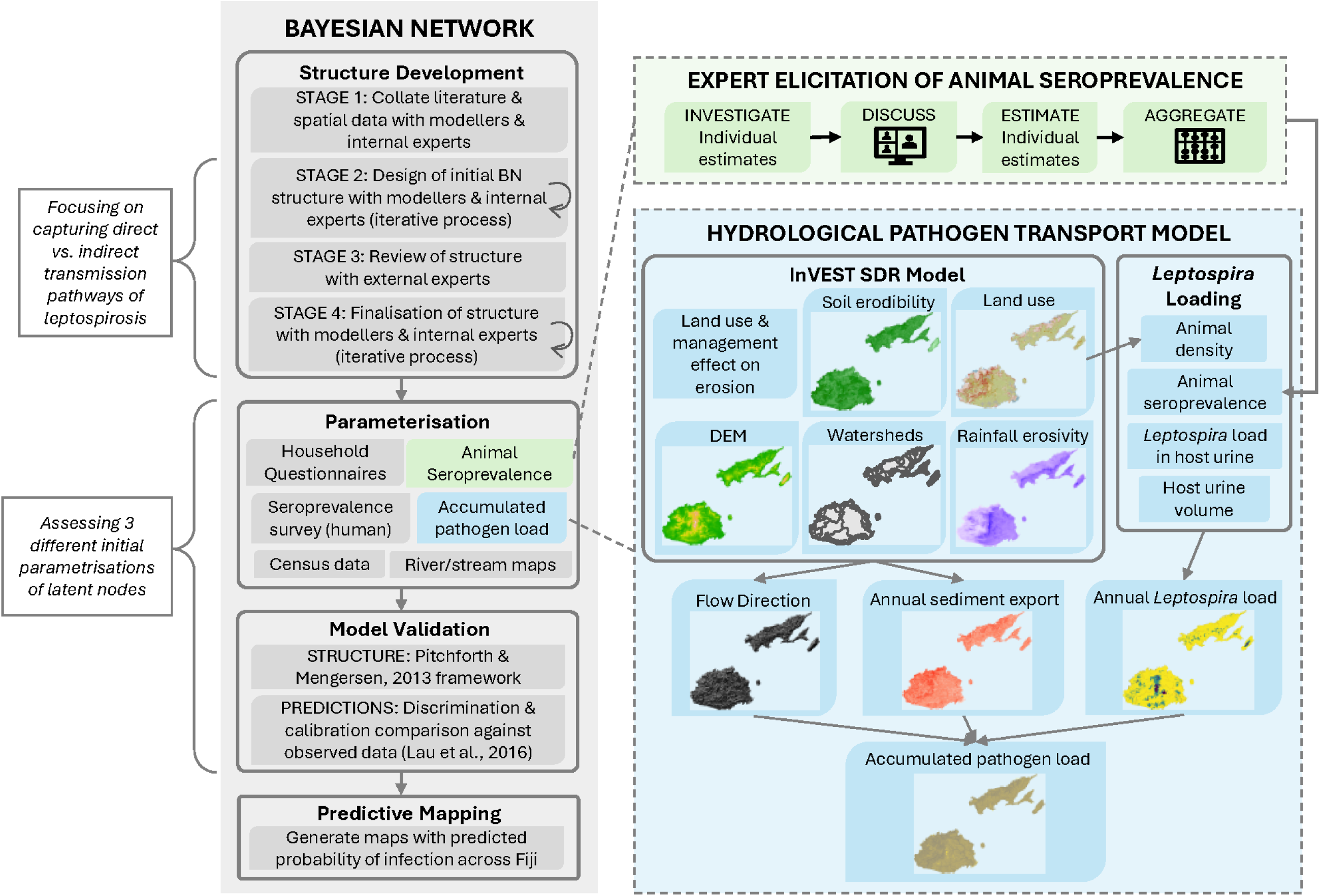
Overview of our hybrid leptospirosis infection model, including the development of the Bayesian network (further detail in Fig 2) and hydrological pathogen transport model. InVEST SDR = Integrated Valuation of Ecosystem Services Sediment Delivery Ratio; DEM = digital elevation model.

### Hydrological pathogen transport model

To develop the hydrological pathogen transport model, we combined estimates of pathogenic *Leptospira* loads entering the environment from key animal hosts with a sediment export model to capture the relative loads of pathogens accumulated across watersheds (Fig 1). Pathogenic *Leptospira* are hypothesised to persist in soil and be transported across environments through soil particles re-suspended during heavy rainfall (8, 10, 11). The hydrological pathogen transport modelling approach captures this accumulation of pathogenic *Leptospira* in the environment as mediated by sediment transport via surface runoff. Using a grid with resolution of three arc-seconds (∼90 m at the equator), the model multiplies an estimate of pathogenic *Leptospira* excreted onto soils by key animal hosts (cells/ha/year) with a modelled estimate of sediment exported to streams via surface runoff (tonnes/ha/year) in each cell across Fiji. The resulting value is then accumulated over land using flow directions determined by a digital elevation model (DEM) before calculating the average accumulated value within a 500 m radius of every pixel (S1 Appendix). The resulting output is a relative indicator/proxy of accumulated pathogenic *Leptospira* load near communities that has been delivered to streams via surface runoff (henceforth called accumulated pathogenic *Leptospira* load).

Pathogen loading within topsoils was estimated by multiplying estimates of seroprevalence, pathogen excretion rates, and population density for each animal species representing an important source of *Leptospira* (e.g., similarly to Barragan et al. (32), Costa et al. (33), and Robinson et al. (34); Fig 1; S2 Appendix). We included only pigs, cattle, and rodents as sources of *Leptospira* as other potential source species (e.g., goats and horses (16)) had limited spatial population data available. We used literature-derived animal occupancy preferences across different land use/cover categories, elevations, and slopes to gain further resolution on the likely distribution of species across Fiji (Table B in S2 Appendix). We derived seroprevalence in pigs, cattle, and rodents based on estimates from four external experts (ethics: 2023/HE000086) using the Investigate, Discuss, Estimate, Aggregate (IDEA) protocol (35), as outlined in S3 Appendix. We used seroprevalence instead of pathogen shedding prevalence (i.e., active infection) based on confidence of experts to provide this information. See Table A and B in S2 Appendix for parameterisation. Soil export to streams was estimated using the widely used sediment delivery ratio model within the Natural Capital Project’s Integrated Valuation of Ecosystem Services and Tradeoffs toolkit (InVEST^®^; version 3.14.2) which uses spatial inputs on environmental data to estimate overland erosion (Fig 1; S5 Appendix). See Table A and B in S4 Appendix for parameterisation.

To calibrate our modelled estimates of accumulated pathogenic *Leptospira* load, we conducted a sensitivity analysis comparing alternative parameterisation of uncertain model assumptions (S5 Appendix). The final hybrid model (combining the pathogen transport model with the BN) used the parameterisation settings which generated pathogen load values most strongly associated with observed leptospirosis seropositivity. This association was assessed using the odds ratio of modelled pathogen load to observed seroprevalence from previously published data (15).

### Bayesian network model development

BNs are a probabilistic modelling tool represented by a directed acyclic graph that uses Bayes’ theorem of conditional probability to assess and update the likelihood of an outcome under different scenarios (36). The drivers and output variables are represented as nodes, which are joined by directional links defining the conditional dependence. Conditional probability tables (CPTs) quantify the likelihood of a node being in a certain state for any given combination of parent nodes. Further background on BNs is provided in S6 Appendix.

The structure of the BN was designed using an iterative process that combined data-driven and expert-led approaches (37, 38). We built the model in four stages involving experienced BN modellers working alongside internal experts from the research team and external experts outside of the research team (Fig 1). This iterative, expert-reviewed process was designed to reduce bias and enabled transparent integration of evidence from both experts and the literature (37–39).

In the first stage of BN structure development, we collated evidence from the literature and spatial data on risk factors relevant to leptospirosis seropositivity in Fiji. Spatial data included risk factors for individuals derived from two household surveys across Fiji (15, 28), population level statistics from national censuses (30, 40, 31), and environmental data from global or national databases (41–43). In the second stage, BN modellers and internal experts used the collated literature and spatial data to develop a preliminary model structure capturing key risk factors (input nodes) driving leptospirosis seropositivity (the outcome node). In the third stage, external experts with knowledge about leptospirosis in Fiji and/or other Pacific Islands were identified by internal experts (or the referring external expert) and recruited via email to review the structure. The external experts reviewed the structure during an in-person workshop or during an online meeting to assess if it accurately represented the direct and indirect leptospirosis transmission pathways relevant to Fiji. In the fourth stage, modellers and internal experts incorporated the feedback from external experts to create a final model structure implemented in the BN software Netica version 6.09 (44).

During the second and fourth stages, both direct and indirect transmission processes were considered. Risk factors were included in the model if experts agreed that: (1) there was a strong evidence-based relationship between the risk factor and leptospirosis in Fiji; (2) the risk factors were not spatially homogenous; and (3) there were sufficient data available on the risk factor across Fiji, or it could be effectively estimated by experts. To allow a more causal exploration of how risk factors influence seroprevalence, we included latent variables to summarise important but largely unobserved and/or data limited pathways (39) between the presence of a risk factor(s) and a human becoming infected (e.g., exposure to an infected animal).

### Hybrid model implementation

The final BN model included 37 variables split into two sub-models that represent the causal pathways driving leptospirosis infection: direct transmission and indirect transmission (Fig 2; S1 Table). The value of the outcome node (leptospirosis seropositivity) was determined by confirmation of infection by either of the sub-models (i.e., infection via either direct or indirect transmission). Eighteen nodes had observed or modelled data available to allow their parameterisation (Table 1). The full structure, list of node states, definitions, and justification of inclusion is provided in S1 Table. Due to data or knowledge gaps, several risk factors identified as important by experts and/or within the literature, for example human mobility, were not included in the model (S2 Table).

**Fig 2.**
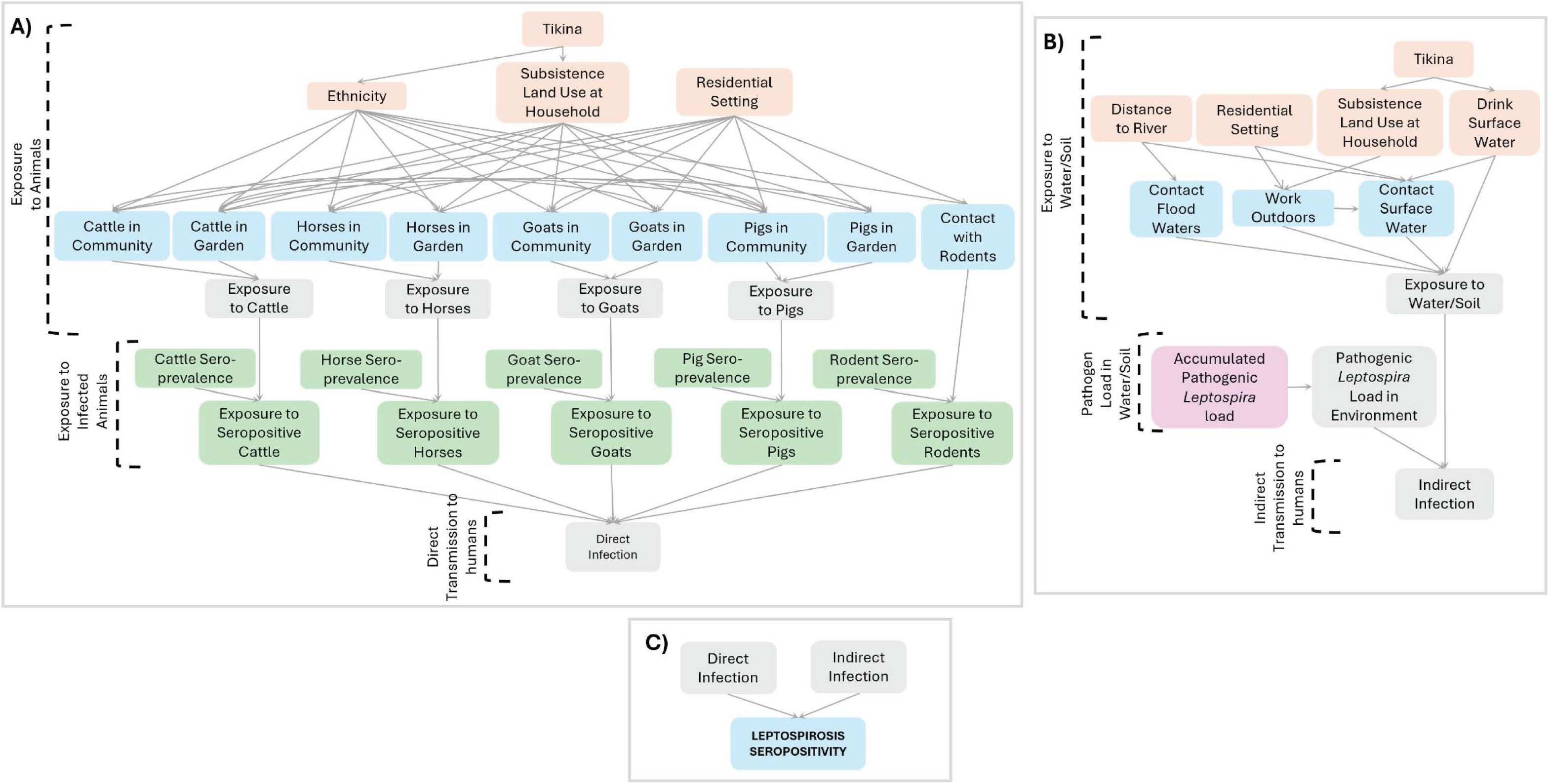
Conceptual diagrams of the Bayesian network predicting leptospirosis seropositivity in Fiji, including the (A) sub-model for direct transmission from animals; (B) sub-model for indirect transmission from the environment; and (C) outcomes of the sub-models used to determine the final outcome node, leptospirosis seropositivity. Node colours represent the data available for the node: orange indicates national census data, green indicates expert elicited data, blue indicates survey data from Lau et al. (15) and/or Jupiter et al. (28), purple nodes indicate data from the hydrological pathogen transport model, and yellow indicates no data (i.e., a latent node). Note: a tikina is a sub-provincial administrative unit in Fiji comprising multiple villages.

**Table 1.** Nodes included in Bayesian network model with data to facilitate their parameterisation. * indicates the node was selected for model calibration validity testing.

| Node(s) | States | Scale of data | Source |
| --- | --- | --- | --- |
| Distance to rivers | Far/Close | 25 m x 25 m river raster | Ministry of Lands and Mineral Resources (45) |
| Residential setting (i.e., rural or urban/peri-urban) | Rural/Urban or Peri-Urban | Tikina-level | Fiji Bureau of Statistics (40) |
|  |  | Data for 2,463 individuals across 111 communities | Lau et al. (15) and Jupiter et al. (28) |
| Ethnicity | Indo-Fijian/<br>*iTaukei*/<br>Other | Tikina-level as % of population | Fiji Bureau of Statistics (30) |
|  |  | Data for 2,463 individuals across 111 communities | Lau et al. (15) and Jupiter et al. (28) |
| Subsistence land use at home | Yes/No | Tikina-level as % of population | Fiji Bureau of Statistics (30) |
| Drinking surface/unprotected ground water at home | Yes/No | Tikina-level as % of population | Fiji Bureau of Statistics (40) |
| Accumulated pathogenic *Leptospira* load * | Very High/<br>High/<br>Medium/<br>Low | 90 m x 90 m raster | Details in S1 Appendix |
| Work outdoors * | Yes/No | Data for 2,463 individuals across 111 communities | Lau et al. (15) and Jupiter et al. (28) |
| Contact surface water * | Yes/No |  |  |
| Contact flood waters * | Yes/No |  |  |
| Cattle/Horses/Goats/Pigs in household garden * | Yes/No | Data for 2,152 individuals across 82 communities | Lau et al. (15) |
| Cattle/Horses/Goats/Pigs in village * | Yes/No |  |  |
| Contact with rodents * | Yes/No |  |  |
| Leptospirosis seropositivity | Yes/No |  |  |

#### Direct transmission

We captured direct transmission through nodes representing an individual’s exposure to key animal hosts, the probability that the animals were infectious, and direct infection from animals to humans (Fig 2A). We were able to include five animal host groups (cattle, horses, goats, pigs, and rodents) within the sub-model. These animals are recognised in the literature and by experts as key drivers of human seroprevalence and have representative data available for human contact or proximity to the animal (e.g., presence within household or community; Table 1). For rodents (i.e., rats and mice), data were available for direct human contact. For livestock animals (i.e., cattle, horses, goats, and pigs), data were available for the presence of animals within the household or community.

To predict human exposure to animals and the risk of subsequent infection, we used socio-demographic data and the expert-derived estimates of the circulation of leptospirosis within animal populations. Socio-demographic data relating to participant ethnicity, subsistence land use at the household, and residential setting (i.e., rural/urban/peri-urban setting) were used to predict human contact with or proximity to the animals (16). We interlinked the presence of different livestock animals within households/communities to account for multicollinearity between variables in the BN (e.g., households who own cows may be more likely to also own pigs) (16). For ethnicity and subsistence land use, we used distributions of the most recent population and housing census results from the Fiji Bureau of Statistics in 2007 and 2017 to help set prior evidence for regions where individual data was not available. Census results were available at the Tikina-level, a sub-provincial administrative unit in Fiji comprising multiple villages (30, 40). To help capture the varying seroprevalence between different animal populations that may influence human infection upon exposure, we included one seroprevalence node for each of the five potential non-human hosts in the model.

#### Indirect transmission

We captured indirect transmission through nodes representing human exposure to surface waters/soils, the load of pathogenic *Leptospira* in those environments, and indirect infection from contaminated environments to humans (Fig 2B). The key routes of human exposure to waters/soils identified by experts and the literature were through working outdoors, contact with flood waters, drinking surface water or unprotected groundwater, and contact from walking, bathing, swimming, or performing household chores (e.g., washing clothes) in surface waters. To predict these exposure routes, we used information on closely associated variables including residential setting, ethnicity, and distance to rivers. Similar to the direct transmission sub-model, we used census data from 2007 and 2017 at the Tikina-level (e.g., ethnicity; (30)) to help set prior evidence for regions without individual data on exposure. We determined the influence of human exposure to contaminated water and/or soil on indirect transmission with a node representing the load of pathogenic *Leptospira* in the environment, which we predict using accumulated pathogenic *Leptospira* load, as developed by our hydrological pathogen transport model.

#### BN parameterisation

We parameterised the BN using a variety of methods depending on the availability of expert knowledge or observed/modelled data (see S1 Table). We first parameterised CPTs where national census results or deterministic rules were available to provide a robust representation of conditions across Fiji. Where possible, we also used expert elicited data to fill CPTs (e.g., for estimates of leptospirosis circulation within animal populations). In cases where data were not known across all of Fiji (i.e., latent nodes and nodes based on household survey data), we used an Expectation Maximisation (EM) algorithm within the Netica software to parameterise CPTs with probabilities that maximize the likelihood of observed/modelled data (46).

To avoid the EM predicting spurious or extreme values for the latent node CPTs, we selected an initial parameterisation (priors) before carrying out the learning to help guide the EM (39). For seven latent nodes, we used a Noisy-OR model to help develop the initial parameterisation (Fig 2; (47, 39)). For these Noisy-OR nodes, we estimated a parameter for each of the parent states, and then the background noise rate to capture unmodelled exposures (see S7 Appendix).

#### Model validation

To assess the influence of initial latent node parameterisation on model outputs, we developed three sets of different initial parameterisation values for latent node priors.

- A non-uniform distribution option where initial parameterisation of latent nodes differentiates the influence of parent nodes (e.g., *Contact with Surface Water* is more likely than *Outdoor Occupation* to cause *Exposure to Water / Soil*);
- A uniform distribution option where initial parameterisation of latent nodes does not differentiate between the influence of parent nodes (e.g., *Contact with Surface Water* is just as likely as *Outdoor Occupation* to cause *Exposure to Water / Soil*); and
- A control without initial parameterisation of latent nodes.

Full details of the initial parameterisation of latent nodes are outlined in S7 Appendix.

We assessed the validity of the three models (developed with either non-uniform, uniform, or control initial parameterisation of latent nodes) using the framework developed by Pitchforth and Mengersen (38). We ensured validity of the model structure and parameterisation through our model development and parameterisation processes wherein we used both the literature and experts to reduce bias and reflect current understanding of the system (S3 Table). To evaluate predictive validity, which addresses confidence in the model behaviour and output, we assessed the discrimination and calibration ability of the three models against observed leptospirosis serological results from Lau et al. (15).

We assessed model discrimination as the ability to correctly predict the outcome variable (i.e., a seropositive result), which we evaluated using the area under the curve (AUC) of the receiver operating characteristic (48). Within Netica, we calculated AUC by randomly assigning 50 % (n = 1,231) of the observed data for parameterising the BN (i.e., training) and tested discrimination of seropositivity against the remaining 50 %. We repeated the process across ten trials to obtain median AUC for each model.

We assessed model calibration as the ability to correctly predict the probability of the outcome given the presence of relevant risk factors, which we evaluated using the relative differences between predicted and observed values and the coefficient of determination (R^2^). We calculated these calibration metrics by training the BNs with the full observed dataset in Netica and comparing the predicted probability of seropositivity against observed seroprevalence when important risk factors were present. We selected 28 risk factor scenarios to assess calibration, representing the states across 13 nodes (Table 1). The risk factors/nodes selected were relevant for potential management projects (e.g., contact with rodents and accumulated pathogenic *Leptospira* load) and had sufficient observed data within our casefile for comparison (i.e., > 100 individuals per state). Nodes that were not considered as an intervention point for environmental management (e.g., ethnicity), or had insufficient observed data (e.g., latent nodes) were not included for calibration (S1 Table).

#### Predictive maps

Using the model with the highest AUC and R^2^, we developed three predictive maps of Fiji depicting: predicted leptospirosis seroprevalence; likelihood of direct infection from animals; and likelihood of indirect infection from the environment. We used GeoNetica software (49) to incorporate spatial data as inputs for selected nodes in the BN, generating a raster of the predicted probability of a specified state within a node of interest (e.g., probability of state *Yes* within the node *Leptospirosis Seropositivity*). We used spatial data on four nodes (tikina, distance to rivers, residential setting, and accumulated pathogenic *Leptospira* load) to generate raster maps, cropping outputs further than 1.5 km from inhabited areas as identified from population grids (50). For the nodes in the BN without corresponding spatial data, GeoNetica estimates the values based on data from other nodes in the CPT.

## Results

Eleven external experts participated in reviewing the model structure. These included experts from government (Fiji Ministry of Agriculture, Fiji Ministry of Health and Medical Services, Ministry of iTaukei Affairs, Department of Environment), regional organizations (Public Health division of the Pacific Community, Live and Learn Fiji, World Wildlife Fund), and academia (from veterinary public health, human public health and environmental health sectors). Four animal health experts from Fiji Ministry of Agriculture, Public Health division of the Pacific Community, and Veterinary Public Health sectors took part in online workshops to estimate leptospirosis circulation within animal populations.

### Hydrological pathogen transport model

Model estimates of the accumulated pathogenic *Leptospira* load proxy were relatively consistent across assumptions tested within our sensitivity analysis, with loads varying across Fiji (Fig B and Table A in S5 Appendix). For the model used as input into the BN, values ranged across 14 orders of magnitude (Fig 3). Predicted loads were generally highest in the water within rivers and on the land immediately adjacent to large rivers, for example the Rewa River flowing through Nausori and Sigatoka River flowing to Sigatoka (Fig 3). This spatial pattern corresponds with the predicted accumulation of pathogen loads over the large catchments flowing into these rivers.

**Fig 3.**
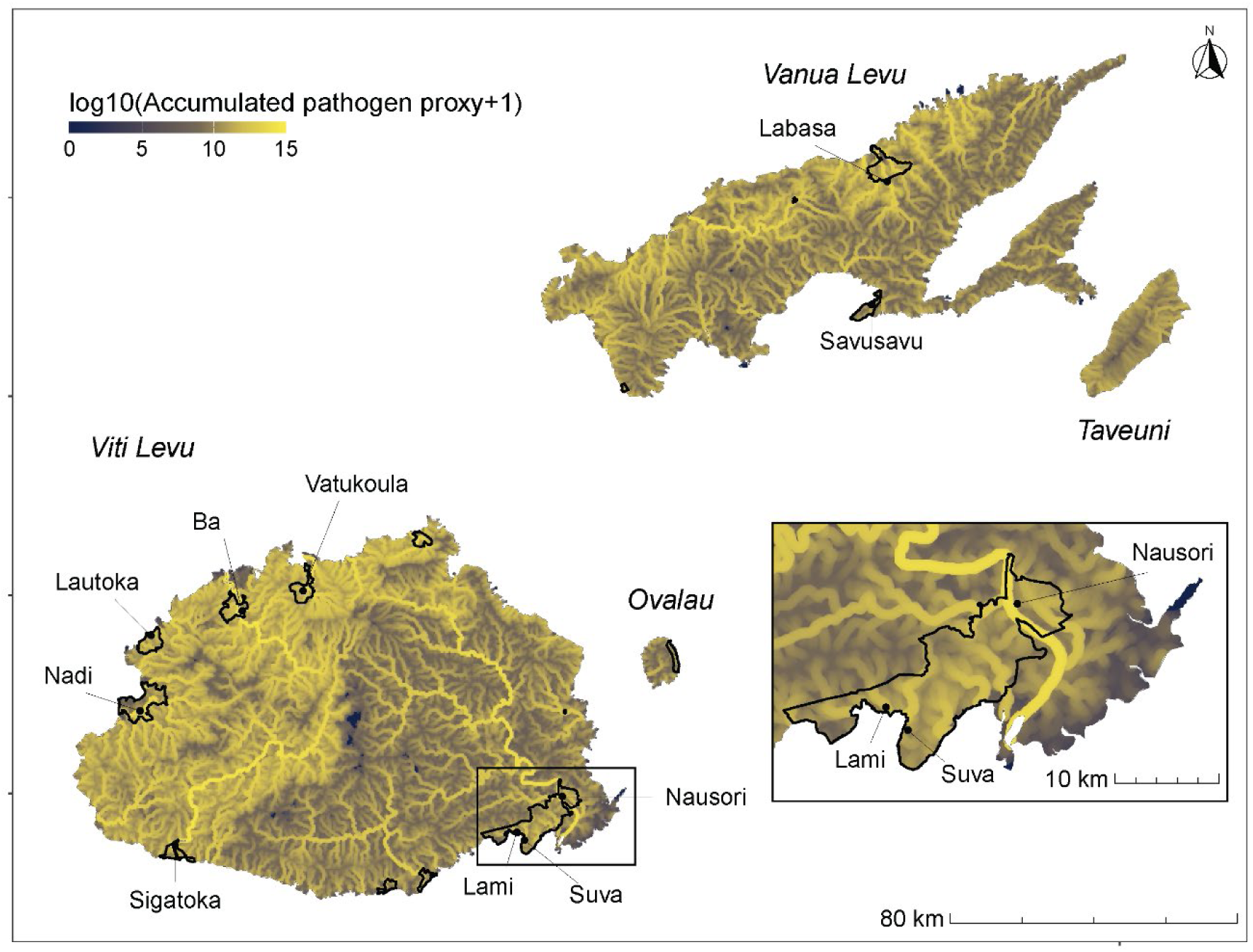
Log transformed accumulated pathogenic Leptospira load (i.e., pathogen load) across four Fijian islands as predicted by the hydrologically linked pathogen model. Urban/peri-urban boundaries and major towns/cities shown in black, and islands labelled in italic text.

Outside of large rivers, pathogen load variation on land mainly corresponded with estimated sediment export (Fig A in S5 Appendix), with the highest accumulated pathogen loads estimated across the northern coast of Vanua Levu and the western half of Viti Levu (Fig 3).

Overall, the spatial pattern of the accumulated pathogen load proxy aligns with known areas of catchment degradation and high sediment export (51–53). Minimal accumulated pathogen loads were predicted within the central highlands of Viti Levu and small coastal peninsulas, corresponding to regions with little accumulation of pathogen (e.g., due to small catchment areas), minimal erosion of sediments (e.g., due to high forest cover), and limited animal host populations (e.g., land use/cover category not suitable for cattle, pigs, or rodents). Discretised values used as input into the BN are given in Fig C in S5 Appendix.

### Hybrid model results

The three hybrid models developed using the same structure, but with either non-uniform, uniform, or control initial parameterisation of latent nodes, produced similar predicted probabilities of national level leptospirosis seropositivity (19.5 %, 20.4 %, and 18.5 %, respectively) when no prior information was provided. All were also close to observed national levels of 19.4 % seroprevalence (Lau et al., 2016; S4 Table). When comparing the likelihoods of transmission routes, there was a slightly higher likelihood of direct infection from animals (11.5 % - 12.6 % likelihood across models) than indirect infection from the environment (7.9 % - 9.1 % likelihood across models) predicted under all models (S4 Table).

The AUC results showed minimal variation over the ten trials and for the three models (S4 Table), with results ranging from 0.60 to 0.65. These AUC results indicate a poor ability to predict an individual’s seropositivity (i.e., discrimination), regardless of which initial parameterisation of latent nodes were used. Model calibration varied more between models, with the non-uniform model performing the best (R^2^ 0.91) when comparing modelled predictions of seroprevalence against average observed seroprevalence under selected risk factor scenarios (Fig 4).

**Fig 4.**
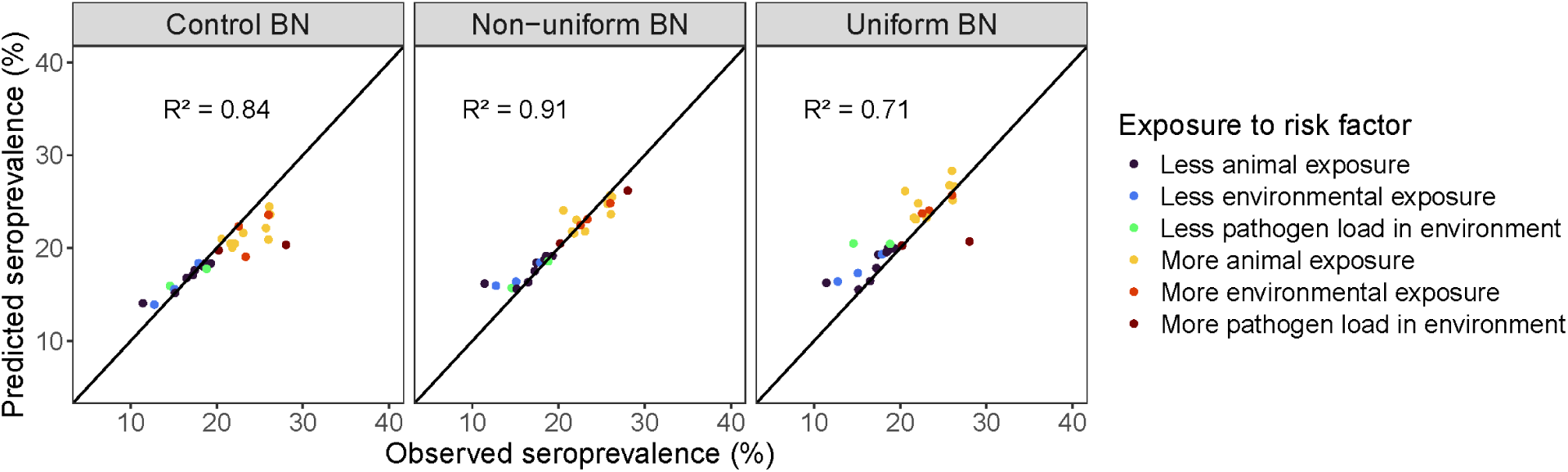
Comparison of model calibration capability between the non-uniform, uniform, and control Bayesian networks (BN). Points show observed vs predicted seroprevalence of individuals under more/less exposure to important risk factors values, and the colour represents the different exposures. Note: more pathogen load in the environment = “High” or “Very High” accumulated pathogenic Leptospira load; less pathogen load in the environment = “Medium” or “Low” accumulated pathogenic Leptospira load.

Across calibration testing, both control and non-uniform BNs consistently predicted the correct direction of influence of risk factors on seropositivity, i.e., predicted values were higher or lower than average seroprevalence (19.4 %) in correspondence with observed values for the scenario (S1 Fig). However, the control model had larger deviations from observed values, resulting in a lower calibration performance (R^2^ 0.84; Fig 4; S1 Fig). In contrast, the uniform model had the worst calibration performance (R^2^ 0.71) due to largely overestimating seroprevalence under the different risk factor scenarios. For example, the uniform model predicted seroprevalence above 19.4% even for lower exposure scenarios such as having low accumulated pathogenic *Leptospira* load, and no goats, horses, pigs, or cows in the household garden (Fig 4; S1 Fig).

### Predictive mapping

The BN with non-uniform priors for the latent nodes (selected as the best performing model) predicted probabilities of leptospirosis seropositivity ranging between 11.0 and 36.8 % across Fiji when provided with spatial inputs (Fig 5A). Urban/peri-urban areas generally had lower predicted probability of seropositivity (mean of 14.8 %) compared to rural areas (mean of 22.4 %), except for urban/peri-urban areas close to rivers, which contained some of the highest predicted probabilities of seropositivity.

**Fig 5.**
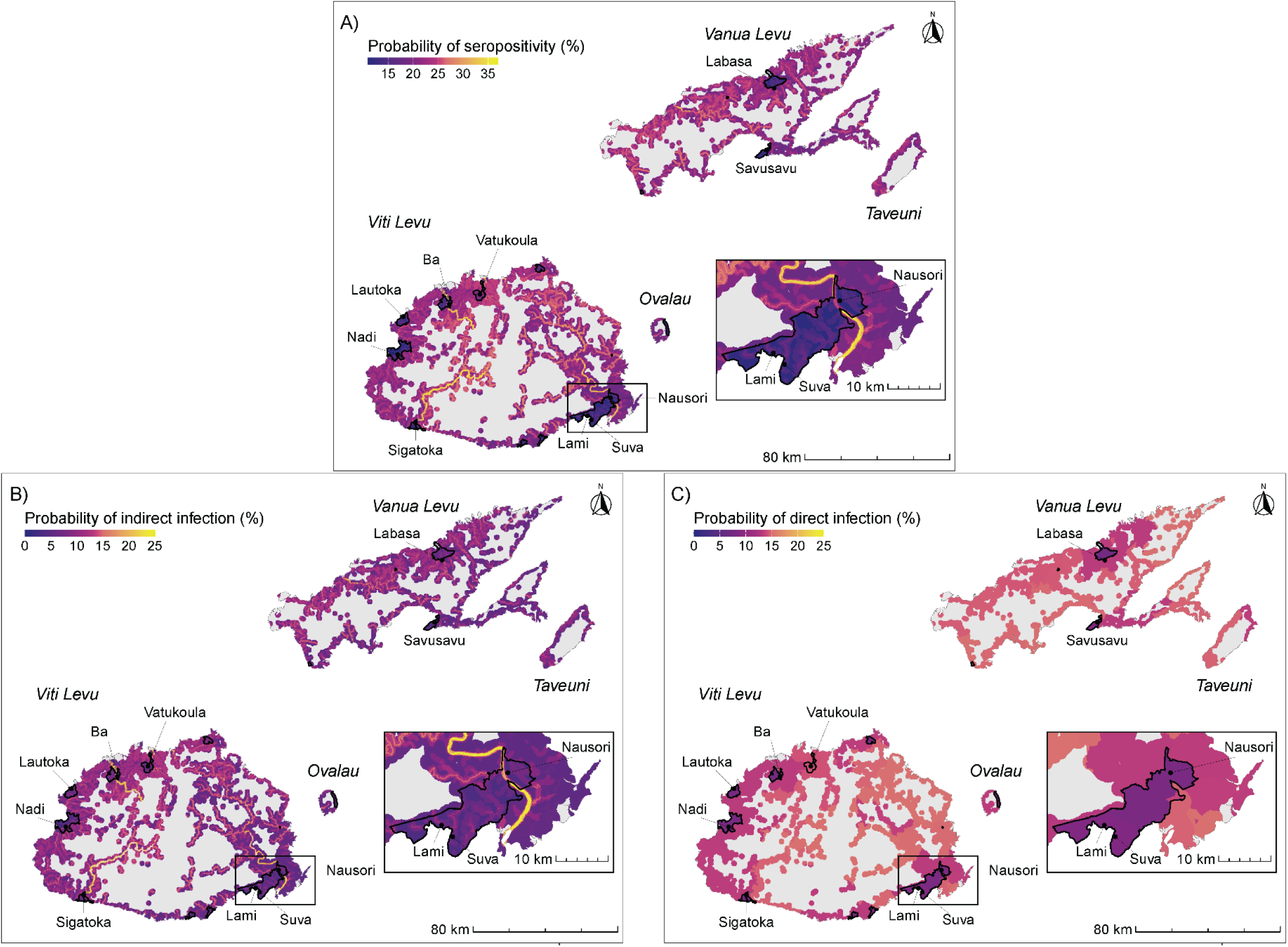
Probability (%) of (A) leptospirosis seropositivity; (B) indirect leptospirosis infection from the environment; and (C) direct leptospirosis infection from animals across four major islands in Fiji (Viti Levu, Vanua Levu, Taveuni, and Ovalau) as predicted by the non-uniform model. Urban/peri-urban boundaries and major towns/cities shown in black, and islands labelled in italic text.

The non-uniform BN predicted substantial spatial variation in the probability of indirect infection from the environment across Fiji, ranging from 3.3 to 25.5 % (Fig 5B). Urban/peri-urban areas generally had lower predicted probability of indirect infection than did rural areas (mean probability of 6.6 % in urban/peri-urban areas vs 9.5 % in rural areas), but the ranges in values were similar (3.3 – 25.5 %), with high probabilities up to 24.7 % still observed in urban/peri-urban areas close to rivers and where accumulated pathogenic *Leptospira* load was high or very high. For example, areas close to the Rewa River, Fiji’s largest river that runs adjacent to Nausori, had generally high predicted probability of indirect infection regardless of urban/peri-urban or rural setting (Fig 5B). The highest probabilities of indirect infection were mostly in the centre west parts of Viti Levu (i.e., within the Sigatoka River catchment; average seroprevalence of 24.7 %), the Rewa River region, and the northern coast of Vanua Levu (Fig 5B), closely matching the distribution of levels of accumulated pathogenic *Leptospira* loads (Fig 3).

Compared to indirect infection, there was less spatial variation in the probability of direct infection from animals, with probabilities ranging between 7.6 to 15.1 % across Fiji (Fig 5C). Urban and peri-urban areas typically had the lowest predicted probabilities of direct infection from animals, such as the highly urbanised area around the country’s capital, Suva, on Viti Levu, and the area around the large towns of Labasa and Savusavu on Vanua Levu. The only exception was the peri-urban area around Vatukoula on Viti Levu, which had one of the highest predicted likelihoods of direct infection from animals (14.3 %). This region was the only urban/peri-urban area to also have a high proportion of households with subsistence land use.

## Discussion

The hybrid model integrating local and watershed-level risk factors with direct and indirect transmission routes provided novel insights into the varying influence of key drivers of leptospirosis in Fiji. High accumulated pathogenic *Leptospira* loads within local environments were influential in driving the predicted spatial patterns of indirect infection and leptospirosis seroprevalence, which notably aligned with known areas of catchment degradation and high sediment export. These spatial patterns highlight the importance of upstream processes in shaping downstream infections: a key dynamic overlooked by previous epidemiological models. Consideration of these upstream processes is critical for informing several key initiatives that are already being explored and implemented by government and non-government organisations in Fiji (54, 55, 28).

Similar to findings from previous studies (17, 18), rural areas and areas close to rivers were associated with higher seroprevalence than were urban/peri-urban areas, areas far from rivers, and areas along the Coral Coast. However, when upstream variables within the watershed are considered, rivers within large catchments (e.g., Rewa River catchment) and/or degraded catchments with large areas of forest loss or agriculture (e.g., Sigatoka River catchment) also had a strong influence on the spatial variation in seroprevalence. This emphasises the importance of taking a One Health approach to managing leptospirosis by demonstrating how socio-demographic and environmental risk factors clearly influence the spatial distributions of infections.

The inclusion of hydrological pathogen transport expands upon existing spatial predictions of leptospirosis infections by capturing the influence of large-scale pathogen transport and notably, allows the assessment of novel and/or large-scale management projects that impact watersheds. Measuring environmental pathogen load across large scales is typically expensive and challenging to model (56), and few field studies have investigated the presence of pathogenic *Leptospira* in open environments (10, 57, 58). Watershed management projects to reduce the spread of pathogens and indirect exposures downstream (e.g., land use planning, erosion control, and drainage improvements) have also had limited evaluation, making it difficult to assess their effectiveness and justify resource allocation for disease prevention and control (4, 5, 8, 28). Using a hybrid modelling approach, we show how these challenges and gaps can be overcome by using the results of a hydrological pathogen transport model to inform an infection model.

Our model was able to identify high-leverage zones where environmental management projects targeting the spread and survival of pathogens (e.g., erosion control) could potentially provide reductions in indirect transmission, such as within the Sigatoka River catchment. As such, while real-world assessments of project impacts are still needed, our model outputs can guide the design of such projects and their evaluation. The use of commonly available datasets, coarse animal population estimates, and a commonly used sediment export model to develop our pathogen transport model also makes this approach more accessible than other more complex pathogen transport models (56).

At a national level, the model predicted a slightly higher likelihood of direct infection from animals than indirect infection from the environment (a difference of 2.4 percentage points). This result is counter to general understanding that indirect transmission is a primary driver of infections in lower and middle-income countries, such as Fiji (7, 4). It is likely that this difference is a result of limited livestock-contact data used in our model, which may have inflated modelled estimates of direct exposure to animal hosts relative to true population-level dynamics. For example, use of personal protective equipment or a person’s age could reduce the risk of direct infection even when animals are present within the household garden or community (9, 4). Furthermore, several risk factors predicting direct transmission are linked with indirect transmission, such as the presence of livestock in the direct transmission sub-model likely correlating with contamination of local environments. As such, the direct transmission sub-model may be capturing additional infection transmission outside of that being directly from the animals themselves, inflating its importance. Further research into transmission pathways is needed to better understand how these factors may influence the relative difference between modes of transmission, as the high levels of subsistence farming and livestock ownership in Fiji (40, 59) may mean that direct transmission from animals is relatively high.

The core objective of the causal modelling approach used here is to allow intuitive predictions of complex disease transmission through the testing of risk factor scenarios, facilitating decision-making on potential disease control strategies. The BN model that applied non-uniform priors was able to reliably capture the influence of these risk factors, as indicated by the summary statistics for model calibration (R^2^ = 0.91). Additionally, we found predictive validity improved when the varying influence of key risk factors was considered within the model, illustrating the importance of capturing complex disease transmission processes. However, our model had limited ability to correctly predict seropositive cases, similar to those published previously for Fiji, including semi-structured BNs (median AUC = 0.59-0.6; (16)), a geographically weighted regression model (mean AUC = 0.64), and a linear regression model (mean AUC = 0.64;(17)). These results were not unexpected given the model’s focus on capturing spatial variation of infection risks rather than diagnosing individual infections.

The relative simplicity of our approach in modelling pathogen transport compared to other pathogen transport models (56) brings about several key limitations. Our approach does not take into account many of the complex processes that determine how *Leptospira* survive within hosts, enter environments, and survive. For example, we do not capture the infection dynamics within and between animal reservoir populations, the varying pathogenicity and environmental survival capability of different *Leptospira* strains, or instream pathogen survival processes (e.g., deposition of sediments or pathogens within rivers). Additionally, we estimated *Leptospira* excreted by animals using seroprevalence in lieu of prevalence of shedding. Limited knowledge of such *Leptospira* dynamics currently prohibits a detailed pathogen transport model from being developed (8, 10), as have been created for other pathogenic bacteria, such as *Escherichia coli* (56). However, recent research predicting pathogenic *Leptospira* loads has found animal abundance to be more important than other factors, such as shedding (60). This supports our pathogen loading modelling approach which uses animal population estimates to predict spatial variation in relative pathogen loading.

Upstream processes are a key driver of leptospirosis risk for those populations living downstream, that until now have remained relatively unexplored. A One Health hybrid modelling approach allows these risks to be considered and can be extended to support implementation of environment-based disease prevention and management strategies. This is a crucial step for water-related infectious disease management and reducing the burden of leptospirosis in Fiji and the Pacific region, and could further assist with interdisciplinary watershed management projects that strive to target multiple drivers of ill-health to people and ecosystems (28).

## Ethics statement

An ethics exemption for expert elicitation of data and review of model structures was granted by the Human Research Ethics Committee of The University of Queensland (2023/HE000086). Ethics approvals for Lau et al. (15) household survey data were granted by the Fiji National Research Ethics Review Committee (2013 03), the Human Research Ethics Committee of The University of Queensland (2014000008) and the London School of Hygiene & Tropical Medicine (6344). Ethics approvals for WISH Fiji household survey data were granted from the Fiji National Health Research and Ethics Review Committee (FNHRERC No: 2018.231.CEN), Fiji National University’s College Health Research Ethics Committee (CHRED ID: 009.19), the University of Sydney’s Human Research Ethics Committee (2019/588) and Edith Cowan University’s Human Research Ethics Committee (#2019–00618). Secondary analysis of an anonymised subset of these datasets was used in the present study.

## Data Availability

Our research includes data from Fiji government, non-government partners, and household surveys that cannot be shared publicly without breaching compliance with data sharing agreements and/or protocol approved by research ethics committees.

## Acknowledgements

The authors would like to thank the many participants, communities, Fiji government and non-government partners, and community health workers who contributed to the collection of field data used in this study from the WISH Fiji project and Lau et al. (15). The authors would also like to thank the experts who provided their input for model structure review and expert elicitation, including Dr Ashnita Prasad, Dr Elva Borja, and Dr Julie Collins-Emerson, and an expert from the Public Health Division SPC. The authors would also like to thank the Bayesian Network Modelling Society for arranging feedback sessions at which this work was presented. Manuscript development was supported by grant #53006 from Bloomberg Philanthropies to the Wildlife Conservation Society (WCS) under the Vibrant Oceans Initiative. Additional support for expert elicitation was supported through funding from the Kiwa Initiative (#CZZ2749 02 L via Agence Française de Développement) to WCS. C.K. is supported by an Australian Research Council Future Fellowship (FT200100314). A.Wa. is supported by an Australian Government Research Training Program Scholarship. C.L.L. and H.J.M. are supported by the University of Queensland Health Researcher Accelerator (HERA) program.

## S1 Appendix. Hydrological pathogen transport model

A strongly hypothesized mode of indirect transmission is through infected animals contaminating water resources and surface soils with their urine, and these contaminated waters and soils moving across lands and downstream into more environments (e.g., water resources, soils) used by humans (8, 10, 11). Livestock and rodents in particular are likely to contribute significant pathogen loads to water resources due to the size and density of livestock and the concentration of *Leptospira* shed by rodents (32, 61). In addition to upstream animals, surface soils are hypothesised to enhance *Leptospira* survival and spread in surface runoff (8, 58, 62), contributing to indirect disease transmission (10). Soils can provide suitable physiological characteristics for *Leptospira* survival in the environment by providing a source of nutrients, accumulating pathogens via adsorption, retaining water, and can enhance spread in the environment through resuspension and runoff (8, 10). Soils that are likely to be important sources of *Leptospira* and facilitate its spread across watersheds are textured clay soils with significant organic matter content, and river soils which retain moisture (8, 58, 62). As such, to help capture indirect transmission in our BN, we developed a hydrological pathogen transport model to estimate the accumulated pathogenic *Leptospira* load in environments near communities. Given strong evidence suggesting topsoil and surface runoff as the main mode of *Leptospira* survival and transport (8, 10), and limited information on possible subsurface transport (58), we focused on representing the transport of sediment-bound pathogens in our model.

In summary, our approach uses modelled annual soil export to streams in tonnes/ha/year (*E*), estimates of pathogenic *Leptospira* in soils in N/ha/year (*PS*), and flow direction as determined by a digital elevation model (DEM), to calculate a unitless value/proxy representing the relative pathogenic *Leptospira* load accumulated across Fiji. Working at the resolution of our DEM (3 arc-seconds, ∼90m at the equator), we calculate the accumulated pathogenic *Leptospira* load (*PW*) in every pixel (*i*) using:

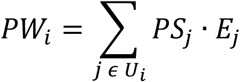

Where *U_i_* are the pixels upstream of stream pixel *i* as determined by flow direction. We then calculate the average accumulated load within a 500m radius of every pixel across Fiji (Fig A). To create relevant states for entry into the BN that represent both the range of values and capture the likely disproportionate influence of larger magnitudes, we discretised the accumulated pathogenic *Leptospira* load into four states based on 50th, 75th, and 99th quantiles.

**Fig A.**
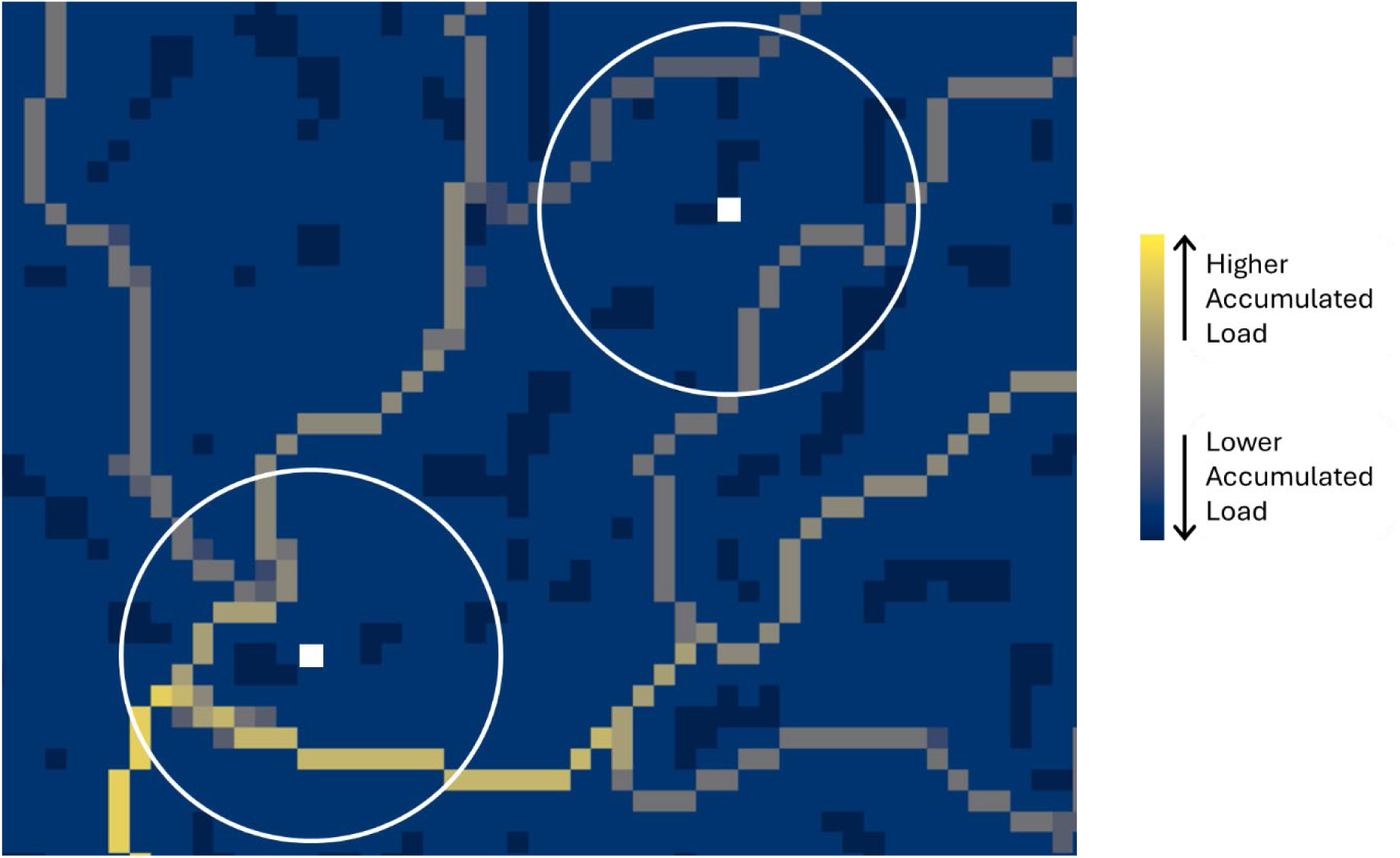
Conceptual approach used to calculate the proxy of accumulated pathogen load near communities from a raster of accumulated pathogen load. We used a 500m search radius around each pixel to calculate the average accumulated load across Fiji. The colour scale represents the accumulated pathogen load, the white pixels represent a hypothetical community pixel of interest for which we want to calculate our proxy of accumulated pathogen load near communities, and the white circles represent the 500m search radius.

## S2 Appendix. Pathogen load from upstream

For input into the hydrological pathogen transport model, we estimate pathogenic *Leptospira* in soils (*PS*) within each pixel *i* using a simple multiplicative model adapted from Barragan et al. (32) and Costa et al. (33):

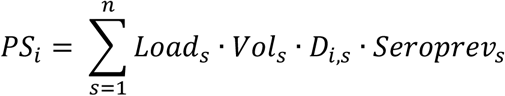

Where for each animal group (*s*), we take the product of the seroprevalence of leptospirosis in their population (*Seroprev*), the average number of *Leptospira* cells excreted per litre of urine when an individual is infected (*Load*), the average litres of urine excreted per individual per day (*Vol*), and the population density in hectares per pixel (*D_i_*_,*s*_). We focus on pigs, cattle, and rodents here, as cattle and rodents have been identified as capable of shedding large amounts of *Leptospira* when infected (32). Additionally, the presence of piggeries upstream of communities has been identified as an important risk factor in other Pacific Islands (61). Other large livestock animals (e.g., goats, horses) may also be important in contributing *Leptospira* to the environment, but data on their populations across Fiji is limited to scales that provide little discriminatory information (e.g., populations only available at the Province level). Expert elicited values for *Seroprev* and literature values for *Load and Vol* per animal group are summarised in Table A.

**Table A.**
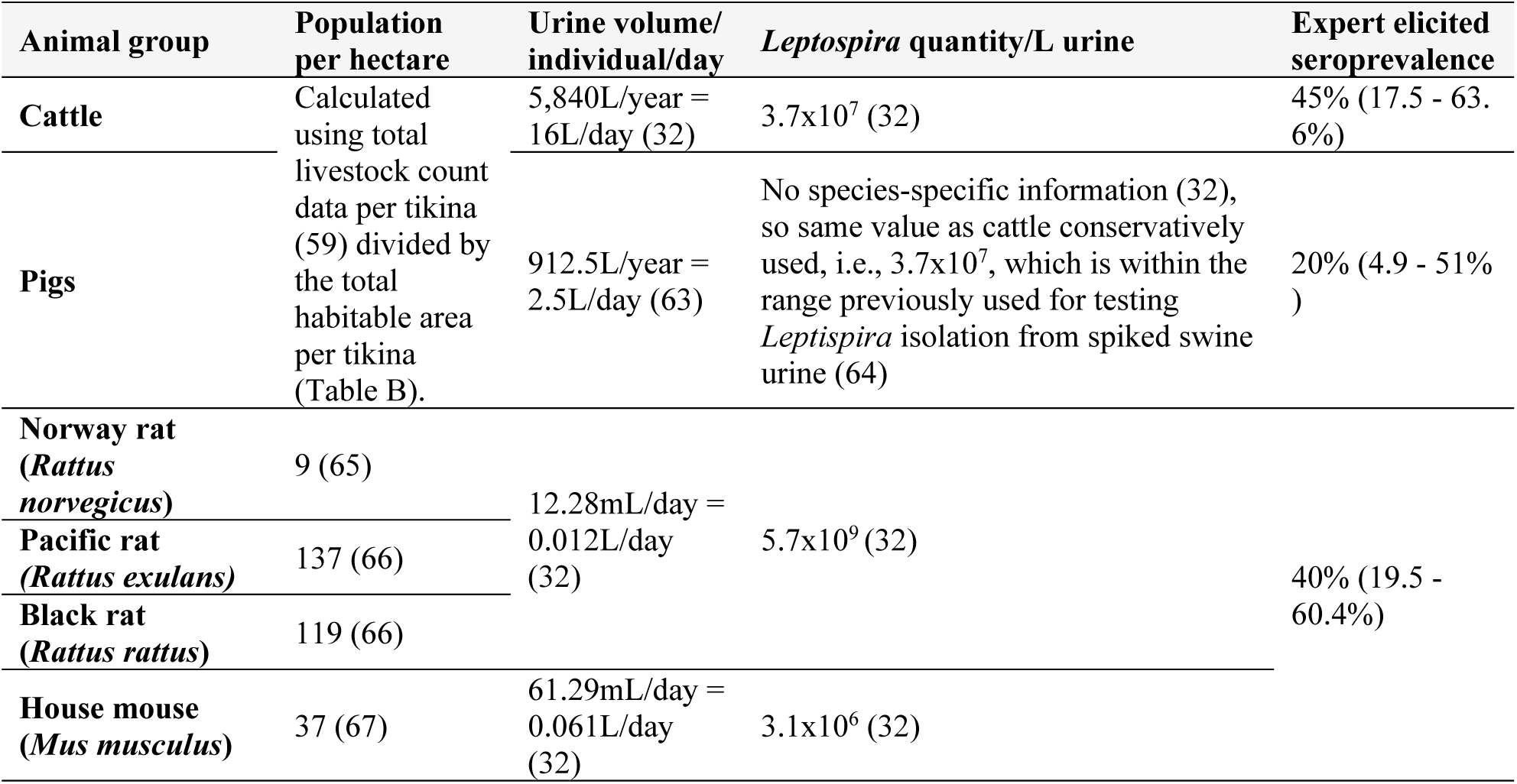
The expert elicited seroprevalence estimates (average best estimate and low/high limits standardised to 80%) and literature-derived pathogen concentration/load and urine volume values used to calculate daily pathogen contribution for each animal group

We used stock counts of cattle and pigs in each tikina/district from the 2009 agricultural census (59) to estimate *D_i_* across Fiji. Due to there being limited data on the distribution of rodent populations across Fiji, we conservatively applied the maximum density estimates from previous studies in Fiji or in the Oceania region for four of the major rodent species in Fiji, black rats (*Rattus rattus*), Pacific rats (*Rattus exulans*), Norway rats (*Rattus norvegicus*), and house mice (*Mus musculus*), obtaining separate *D_i_* values for each.

Instead of directly applying rodent densities and cattle populations across all of Fiji, we used information on land use/cover category, elevation, and slope to gain further resolution on the likely distribution of species across Fiji and also better assess the area for calculating *D_i_* for cattle and pigs (Fig A). This approach is similar to one used by another study in Hawaii attempting to estimate the distribution of a key pathogen host species with limited data (34). We excluded all animal groups from water (i.e., set density to 0) and steep slopes (>40 degrees). Steep slopes were excluded to avoid unrealistically high delivery ratios, similarly to Robinson et al. (34), and also reflect the likely resident densities in these areas. We further excluded rodents from remote intact forested areas (>4km from forest edges) where populations are likely to be limited (68). We additionally used distribution information published by the Ministry of Agriculture and Waterways on the land use/cover categories occupied by feral cattle, pigs, as well as black rats, Pacific rats, Norway rats, and house mice (69). For example, Pacific rats were excluded from forest/tree land cover categories at high elevations (>600m elevation) but not low elevation (<600m elevation) as per their described habitat use (69). Feral cattle and pig habitats reported by the Ministry of Agriculture & Waterways were used to assist in assigning livestock distribution as these animals are known to have wide foraging environments in Fiji. For example, even when farmed, livestock can be well outside of pastures/grazing land and include areas such as coconut plantations, trees, and other crops. We then applied the rodent densities from the literature only to areas where rodents are likely to inhabit. For cattle and pigs, we divided the cattle and pig populations by the area they are likely to inhabit per tikina to calculate *D_i_* for each tikina, and then only applied these densities to those areas. Full details of the distribution of animals (i.e., presence/absence) within each land use/cover category is summarised in Table B.

**Table B.** The literature derived presence/absence (1/0) of each animal group within land use/cover categories across Fiji. Sources for determining animal presence/absence in land use/cover categories were Naikatini et al. (69) and Olson et al. (68) for rodents, and Naikatini et al. (69) for livestock.

| Land use/cover category | House mouse<br>(*Mus musculus*) | Pacific rat<br>(*Rattus exulans*) | Black rat<br>(*Rattus rattus*) | Norway rat<br>(*Rattus norvegicus*) | Cattle | Pig |
| --- | --- | --- | --- | --- | --- | --- |
| Water | 0 | 0 | 0 | 0 | 0 | 0 |
| Wetland* | 0 | 1 | 1 | 1 | 0 | 0 |
| Shrubland | 0 | 1 | 1 | 1 | 0 | 1 |
| Mixed cassava/tobacco crops* | 1 | 1 | 1 | 1 | 1 | 1 |
| Grassland* | 0 | 1 | 1 | 1 | 1 | 1 |
| Coconut plantations* | 1 | 1 | 1 | 1 | 1 | 1 |
| Remote native forest* | 0 | 0 | 0 | 0 | 0 | 0 |
| Softwood plantation forest* | 0 | 0 | 1 | 0 | 1 | 1 |
| Hardwood plantation forest* | 0 | 0 | 1 | 0 | 1 | 1 |
| Native forest >600m* | 0 | 0 | 1 | 1 | 0 | 1 |
| Native forest <600m* | 0 | 1 | 1 | 1 | 1 | 1 |
| Other | 0 | 0 | 1 | 0 | 1 | 1 |
| Settlement* | 1 | 1 | 1 | 1 | 1 | 1 |
| Sugarcane/fruits* | 1 | 1 | 1 | 1 | 1 | 1 |
| Mixed Crops | 1 | 1 | 1 | 1 | 1 | 1 |
| Mixed grazing land* | 1 | 1 | 1 | 1 | 1 | 1 |
| Cropland/agriculture | 1 | 1 | 1 | 1 | 1 | 1 |
| Barren Land | 0 | 0 | 1 | 0 | 0 | 0 |
| Bare soil/rock | 0 | 0 | 1 | 0 | 0 | 0 |
| >40 degree slope | 0 | 0 | 0 | 0 | 0 | 0 |
| **\*Land use/cover categories that were combined and/or calculated as below** |  |  |  |  |  |  |
| >40 degree slope | >40 degree slope areas calculated from DEM of Fiji (see Table A in S4 Appendix for DEM details). |  |  |  |  |  |
| Wetland | Swamp vegetation, wetland, mangrove |  |  |  |  |  |
| Mixed cassava/tobacco crops | Cassava, Dalo/Yagona/Cassava/Yam, Tabacco/Cassava/Okra/Bean/Cucumber/Eggplant, Tobacco |  |  |  |  |  |
| Grassland | Open Land/Grassland, grassland |  |  |  |  |  |
| Coconut plantations | Coconut Crops, Coconut Forest, Coconut Plantation, Dalo/Yagona/Coconut |  |  |  |  |  |
| Settlement | Settlement, urban/impervious |  |  |  |  |  |
| Remote native forest* | Tree/forest cover that is >900m elevation (as per DEM) and/or > 4 km from forest edges within native forest patches defined by forest cover map (70) |  |  |  |  |  |
| Softwood plantation forest* | Tree/forest cover that overlaps with softwood (i.e., pine) timber plantation forest defined by forest cover map (70) |  |  |  |  |  |
| Hardwood plantation forest* | Tree/forest cover that overlaps with hardwood (i.e., mahogany) timber plantation forest defined by forest cover map (70) |  |  |  |  |  |
| Native forest >600m | Tree/forest cover that is > 600 m elevation (as per DEM), is outside of remote native forest (defined above), but is within native forest patches defined by forest cover map (70) |  |  |  |  |  |
| Native forest <600m | Tree/forest cover that is < 600 m elevation (as per DEM), is outside of remote native forest (defined above), but is within native forest patches defined by forest cover map (70) |  |  |  |  |  |
| Sugarcane/fruits | Sugarcane, Pineapple/Mango/Banana/Floriculture |  |  |  |  |  |
| Mixed grazing land | Grazing Land / Trace of Maize / Paddy |  |  |  |  |  |
| Wetland | Swamp vegetation, wetland, mangrove |  |  |  |  |  |

## S3 Appendix. Expert elicitation of animal seroprevalence

The relative circulation of leptospirosis within animal populations is an important factor in determining human infections. We used estimates of animal seroprevalence (i.e., the proportion of the animal population with *Leptospira* antibodies in their blood indicating past infection), as inputs for parameterising animal infection nodes within our BN and the *Leptospira* loads in our hydrological pathogen transport model. We use seroprevalence instead of prevalence (i.e., active infection, which corresponds better with pathogen shedding; (71)) given the absence of prevalence data in animal populations in Fiji and/or across the Pacific (61) and because experts were generally uncomfortable providing prevalence estimates for animal groups. While seroprevalence data is also limited in Fiji, seroprevalence was accepted by experts as a suitable proxy of leptospirosis circulation in animals for our purposes, and experts were more comfortable with making seroprevalence estimates given the presence of at least some seroprevalence evidence from across other Pacific Islands (61). As such, we used a structured expert elicitation process to obtain estimates of seroprevalence in key animal reservoir populations in Fiji. Estimates were obtained from four of our five external subject matter experts recruited to review the BN. The selected external experts had experience in leptospirosis infections in Pacific Island animals. Experts included representatives from the Fijian Ministry of Agriculture, veterinary public health research, and The Pacific Community.

We obtained estimates from experts using a robust protocol for structured expert elicitation, namely the “Investigate,” “Discuss,” “Estimate” and “Aggregate” (IDEA) protocol (35). We provided experts with a questionnaire (example questionnaire below) to facilitate the structured elicitation of an individual expert’s best estimates of seroprevalence for each animal group, their lowest and highest estimate (i.e., credible interval), as well as their confidence in their estimates (“Investigate”). Our questionnaire included background information on previous animal seroprevalence surveys in Fiji and across Pacific Islands (61, 72, 73) to help experts with developing their estimates. Experts were then invited to an online group meeting, facilitated by the BN modellers, to discuss the range of individual anonymised estimates with credible intervals standardised to 80% using the reported confidence (“Discuss”) (35). Following the group discussion, experts were able to adjust their estimates based on any new information from the discussion, if desired (“Estimate”), and the final individual expert estimates were aggregated using quantile aggregation (“Aggregate”) (35). Using the package ‘triangle’ in R (74), we created a triangular distribution of seroprevalence from the final estimates for each animal group, from which the probability of seroprevalence within intervals of 10% was calculated (using a sample of 100,000). These probabilities were used for parameterising animal seroprevalence nodes in the BN (described in S1 Table), and the aggregated best estimates were also used to help assess the proxy of accumulated pathogenic *Leptospira* across Fiji as used in the hydrological model (described in S2 Appendix).

### Example expert elicitation questionnaire

Q1: What is the average seroprevalence of pathogenic leptospirosis in **cattle** in Fiji? Clarification:

Cattle have been identified as reservoirs of leptospirosis, and have been associated with human infections across many Pacific Islands (61, 75), including Fiji (15, 16). We were able to identify two seroprevalence estimates from the literature summarised in a recent review (61), however both have limitations in terms of adequately representing current cattle populations across Fiji:

– 27.5% seroprevalence from a study published in **<u>1984</u>** surveying 480 cattle (outdated)
– 77.9% seroprevalence from a study published in 2016 surveying **<u>9</u>** cattle (small sample)

Additional Information:

Published leptospirosis seroprevalence in cattle across the Pacific Islands ranges from 1.4 – 100%, (figure below) when considering results published since the 1980’s (61). This includes 18 seroprevalence estimates.

**Fig.**
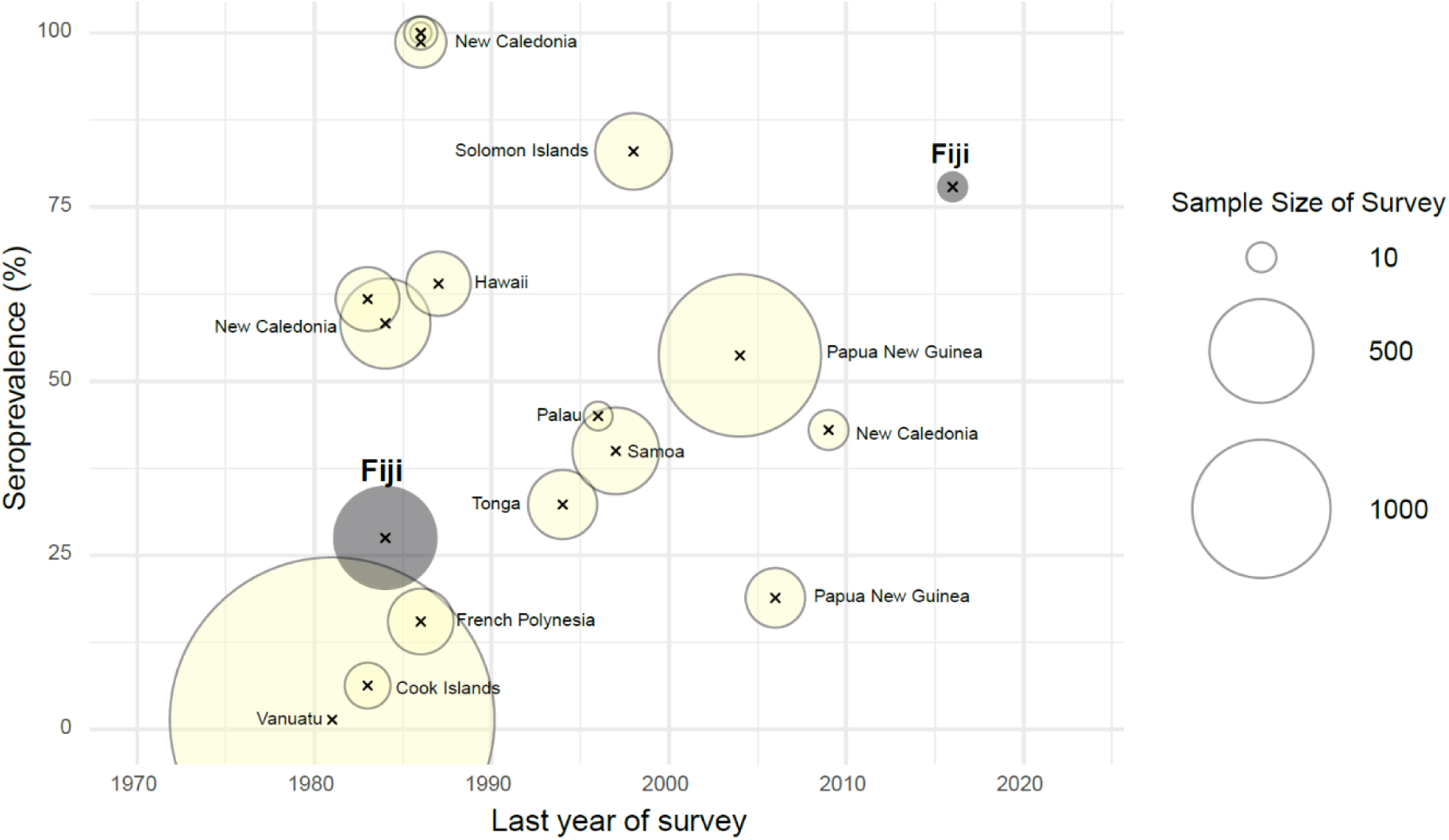
Reported leptospirosis seroprevalence from cattle populations in 11 Pacific Islands (61). Circle size is proportional to survey sample size and samples from Fiji are grey.

Your estimate for Question 1:

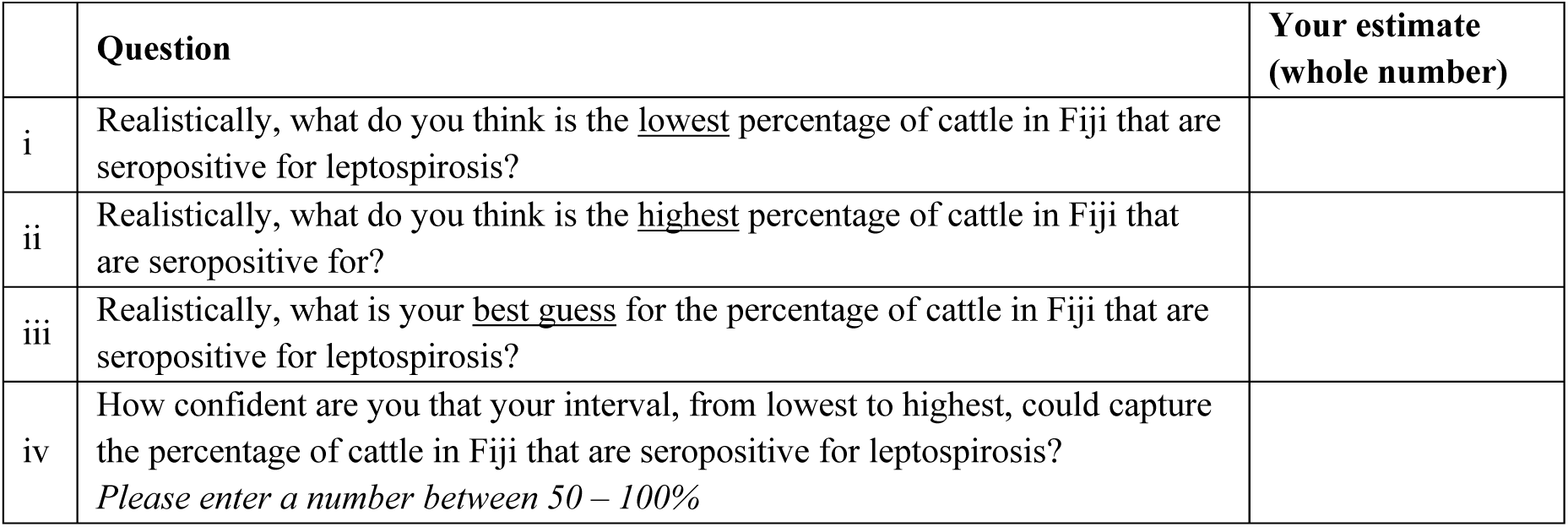

Your Comments:

Please enter any comments, additional knowledge (e.g., data sources, publications) or justification that you have about this question and/or your estimate. This may be shared (anonymously) with the group in Round 2.

## S4 Appendix. Sediment export

We used the sediment delivery ratio (SDR) model within the Natural Capital Project’s InVEST^®^ (Integrated Valuation of Ecosystem Services and Tradeoffs) toolkit (version 3.14.2; (76)) to model annual exported sediment (full description in Hamel et al. (77)). Although InVEST’s SDR model does not capture all erosional processes (e.g., gully erosion or landslides), the minimal parameter requirements and capacity to use globally available data makes it highly accessible for data-poor contexts, such as Fiji (78), and has been shown in other volcanic Pacific Island settings to produce better estimates of sediment export than some more complex approaches when only limited data is available (79).

For use in Fiji, we have parameterised the model in ways shown to better reflect observed sediment yields in other volcanic Pacific Islands (79), outlined in Table A. For cover management factor, we used values found from land use category across extensive research parameterizing SDR models globally, and protection factor was set as the default of 1 (Table B). We developed mosaics of separate rasters for both the DEM and land use due to gaps in the data available (e.g., the raster not covering coastlines or missing data past the antemeridian). All spatial inputs were wrangled and projected to EPSG:3460 within R (74).

**Table A.** Development of inputs and relevant data sources used for parameterising sediment delivery ratio model

| Data | Parametrisation | Source(s) |
| --- | --- | --- |
| **Conditioned digital elevation model (DEM)** | We developed a conditioned DEM raster for Fiji using two DEMs, both of which are sourced from Shuttle Radar Topography Mission data. The globally available conditioned DEM from HydroBASINS (resolution 3 arc-seconds, ~90 m at the equator) was extended past the anti-meridian using a DEM from The Pacific Community (SPC) to ensure full coverage of land across all islands. We then conditioned the mosaic DEM using the *whitebox* package (80, 81) in R to fill sinks, breach depressions (least cost method with search distance of 3 cells), and fill depressions (Wang Liu method). | Conditioned DEM from HydroBASINS (43); DEM from SPC obtained from the Wildlife Conservation Society. |
| **Watershed polygon** | We developed watershed polygons for Fiji from the conditioned DEM (described above) using the *whitebox* package (80, 81) in R and pour points defined as the endpoints of small streams (>5 flow accumulation) to ensure coverage of even the small coastal watersheds. | - |
| **Rainfall erosivity (R factor)** | We developed a rainfall erosivity raster for Fiji from a globally available mean annual rainfall raster at 30 second resolution. We multiplied annual rainfall by 0.5 to calculate rainfall erosivity as per Roose equation (82) and resampled the raster to match the DEM. The Roose equation has been shown to produce sediment export values a magnitude closer to observed ranges compared to other equations (79), and we selected a multiplier of 0.5 as is within the range expected for mountainous tropical areas near the ocean (0.2-0.6) (83) and has also been used successfully in other volcanic Pacific Islands (79, 84). | Rainfall from WorldClim (41) |
| **Soil erodibility (K factor)** | We developed a soil erodibility raster for Fiji from a globally available 1km resolution K factor raster. We filled gaps present at the anti-meridian with the modal value of surrounding cells, using a focal (“moving window”) calculation in R, and resampled the raster to match the DEM. | K<sub>GloSEM</sub> developed by Gupta et al. (42) |
| **Land use/cover (LULC)** | We developed a LULC raster for Fiji using a combination of LULC maps, all of which are available from SPC. We combined the high resolution LULC rasters for all Fijian tikinas from Davies et al. (85) (10x10m), and extended the coverage to include the eastern edge of Vanua Levu using an older, lower resolution LULC map (86) (25x25m) which used a combination of data (e.g., satellite imagery, imagery of vintage resources, agriculture census). We resampled the extended raster to match the DEM. | (85, 86) |
| **Biophysical Table** | This defines the C factor and P factor for each land use/cover category. See Table B for full values. |  |
| **Threshold flow accumulation** | 100 | To ensure small coastal flows were captured |
| **Borselli k Parameter** | 2 | Default values |
| **Max SDR Value** | 0.8 |  |
| **Borselli IC0 Parameter** | 0.5 |  |
| **Max L Value** | 122 |  |

**Table B.** Biophysical table used for sediment delivery ratio model including cover management factor (C factor), support practise factor (P factor), and references attributed to each land use/cover category.

| Code | Land use/cover categories | C factor |  | P factor | Reference |
| --- | --- | --- | --- | --- | --- |
|  |  | min | max |  |  |
| 1 | Water | 0 | 0 | 1 | Falinski (87) |
| 2 | Wetland* | 0.001 | 0.001 | 1 | Wischmeier et al. (83); Falinski (87) |
| 3 | Shrubland | 0.007 | 0.15 | 1 | FAO (88) |
| 4 | Mixed cassava/tobacco crops* | 0.01 | 0.8 | 1 | FAO (88) |
| 5 | Grassland* | 0.01 | 0.4 | 1 | FAO (88) |
| 6 | Coconut plantations* | 0.1 | 0.3 | 1 | FAO (88) |
| 7 | Remote native forest* | 0.001 | 0.006 | 1 | Wischmeier et al. (83); FAO (88) |
| 8 | Softwood plantation forest* | 0.007 | 0.009 | 1 | FAO (88) |
| 9 | Hardwood plantation forest* | 0.01 | 0.05 | 1 | FAO (88); Chicas and Omine (89) |
| 10 | Native forest > 600 m* | 0.001 | 0.006 | 1 | Wischmeier et al. (83); FAO (88) |
| 11 | Native forest < 600 m* | 0.001 | 0.006 | 1 | Wischmeier et al. (83); FAO (88) |
| 12 | Other | 0 | 0 | 1 | Falinski (87) |
| 13 | Settlement* | 0.1 | 0.2 | 1 | Wischmeier et al. (83); FAO (88) |
| 14 | Sugarcane/fruits* | 0.1 | 0.5 | 1 | FAO (88) |
| 15 | Mixed Crops | 0.3 | 0.4 | 1 | FAO (88) |
| 16 | Mixed grazing land* | 0.1 | 0.9 | 1 | FAO (88) |
| 17 | Cropland/agriculture | 0.1 | 0.9 | 1 | Wischmeier et al. (83); FAO (88) |
| 18 | Barren Land | 0.85 | 0.85 | 1 | Roose (90) |
| 19 | Bare soil/rock | 1 | 1 | 1 | FAO (88) |
| **\*Land use/cover categories that were combined across categories or calculated** |  |  |  |  |  |
|  | **Wetland** | Swamp vegetation, wetland, mangrove |  |  |  |
|  | **Mixed cassava/tobacco crops** | Cassava, Dalo/Yagona/Cassava/Yam, Tabacco/Cassava/Okra/Bean/Cucumber/Eggplant, Tobacco |  |  |  |
|  | **Grassland** | Open Land/Grassland, grassland |  |  |  |
|  | **Coconut plantations** | Coconut Crops, Coconut Forest, Coconut Plantation, Dalo/Yagona/Coconut |  |  |  |
|  | **Settlement** | Settlement, urban/impervious |  |  |  |
|  | **Remote native/cloud forest\*** | Tree/forest cover that is > 900 m elevation (as per DEM) and/or > 4 km from forest edges within native forest patches defined by forest cover map (70) |  |  |  |
|  | **Softwood plantation forest\*** | Tree/forest cover that overlaps with softwood (i.e., pine) timber plantation forest defined by forest cover map (70) |  |  |  |
|  | **Hardwood plantation forest\*** | Tree/forest cover that overlaps with hardwood (i.e., mahogany) timber plantation forest defined by forest cover map (70) |  |  |  |
|  | **Native forest > 600 m** | Tree/forest cover that is > 600 m elevation (as per DEM), is outside of remote native/cloud forest (defined above), but is within native forest patches defined by forest cover map (70) |  |  |  |
|  | **Native forest < 600 m** | Tree/forest cover that is < 600 m elevation (as per DEM), is outside of remote native/cloud forest (defined above), but is within native forest patches defined by forest cover map (70) |  |  |  |
|  | **Sugarcane/fruits** | Sugarcane, Pineapple/Mango/Banana/Floriculture |  |  |  |
|  | **Mixed grazing land** | Grazing Land / Trace of Maize / Paddy |  |  |  |

## S5 Appendix. Assessing hydrological model assumptions

Given the lack of data to help validate the accumulated pathogenic *Leptospira* load values for Fiji, we conducted a sensitivity analysis to help calibrate model parameters. We compared two methods for developing sediment export estimates and two methods for developing pathogen in topsoil estimates to assess how they might influence predictions of leptospirosis seropositivity. We vary the cover-management factor (C factor) associated with each land use/cover category in the sediment export model to generate low and high estimates of sediment export for comparison (Table B in S4 Appendix). C factor is one of the most sensitive parameters used within the SDR model (91), and is also often the most relevant for assessing environmental policy and land use as it represents each land use/cover category’s propensity to erode soil due to factors which can be altered by human intervention (e.g., vegetation cover and soil management). To assess the influence of these land use distribution estimates, we also create two pathogen load estimates across Fiji, with and without modifying animal density by land use. This resulted in four alternative estimates of modelled accumulated pathogenic *Leptospira* load:

- Low C factor, animal density varied by land use
- High C factor, animal density varied by land use
- Low C factor, animal density not varied by land use
- High C factor, animal density not varied by land use

As expected, we find sediment export per watershed varies drastically between low and high C factor estimates, however the relative differences between watersheds are similar (Fig A). Also as expected, the relative distribution of pathogen load excreted into topsoil by animals changed when animal density considered land use/cover (Fig A in Appendix). The changes in these assumptions created some variation in the final accumulated pathogen load values calculated across Fiji, with high value areas staying relative constant, and more variation seen in areas modelled as having low values of the proxy (Fig B).

We assessed the association between the final accumulated pathogen load values and leptospirosis seropositivity through simple univariate logistic regressions (using the *‘lme4’* package in R (74)), and using seropositivity data from 2,152 individuals as reported in Lau et al. (15). All four estimates were statistically significant, but we found the accumulated pathogen load developed with high C factor estimates for sediment and variable animal density provided a more informative risk (Table A), and so was selected for use in the BN (Table B; Fig C).

**Fig A.**
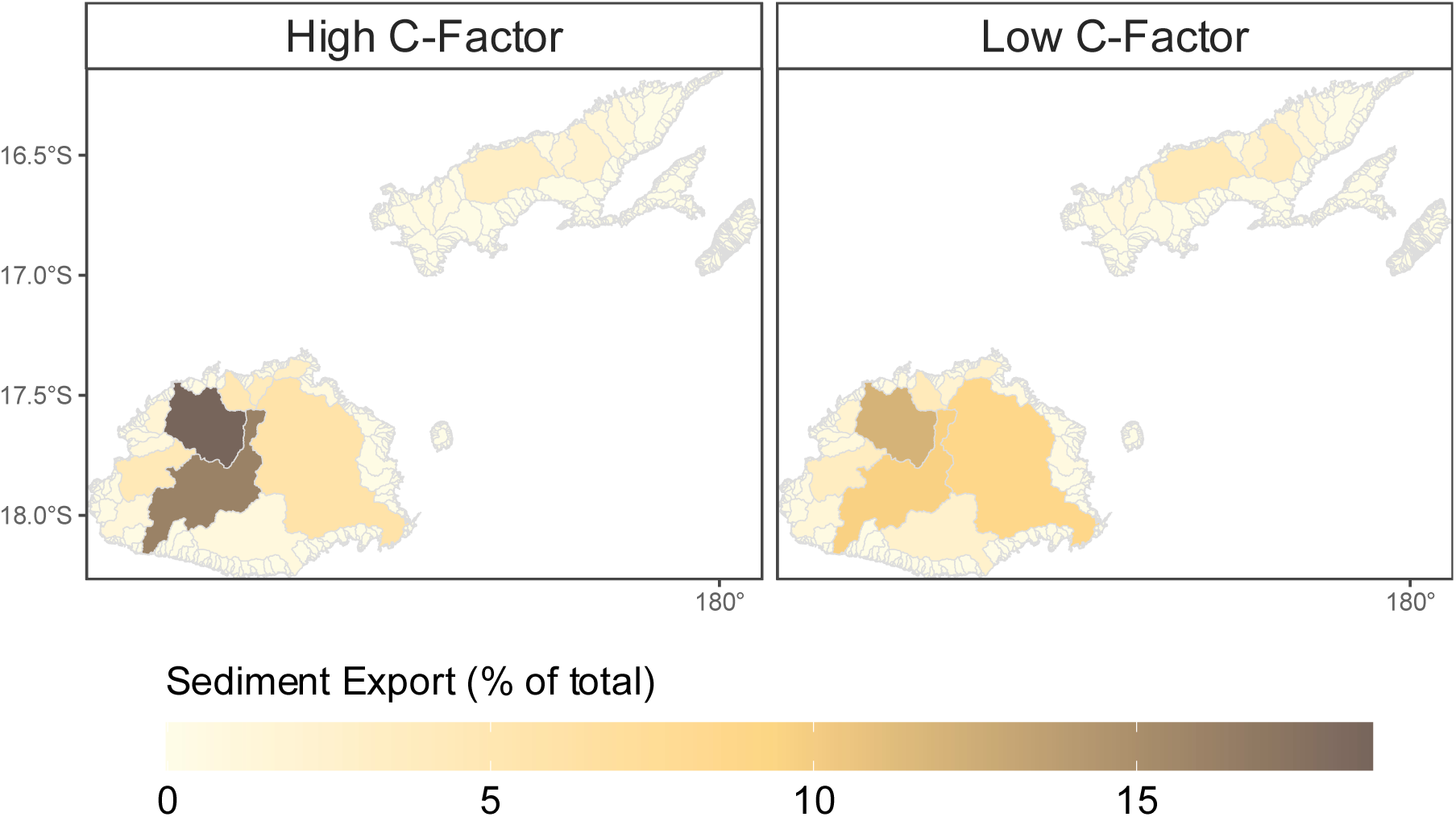
Sediment export within watersheds across Fiji as estimates by InVEST SDR model using high C factor estimates (A) and low C factor estimates (B).

**Fig B.**
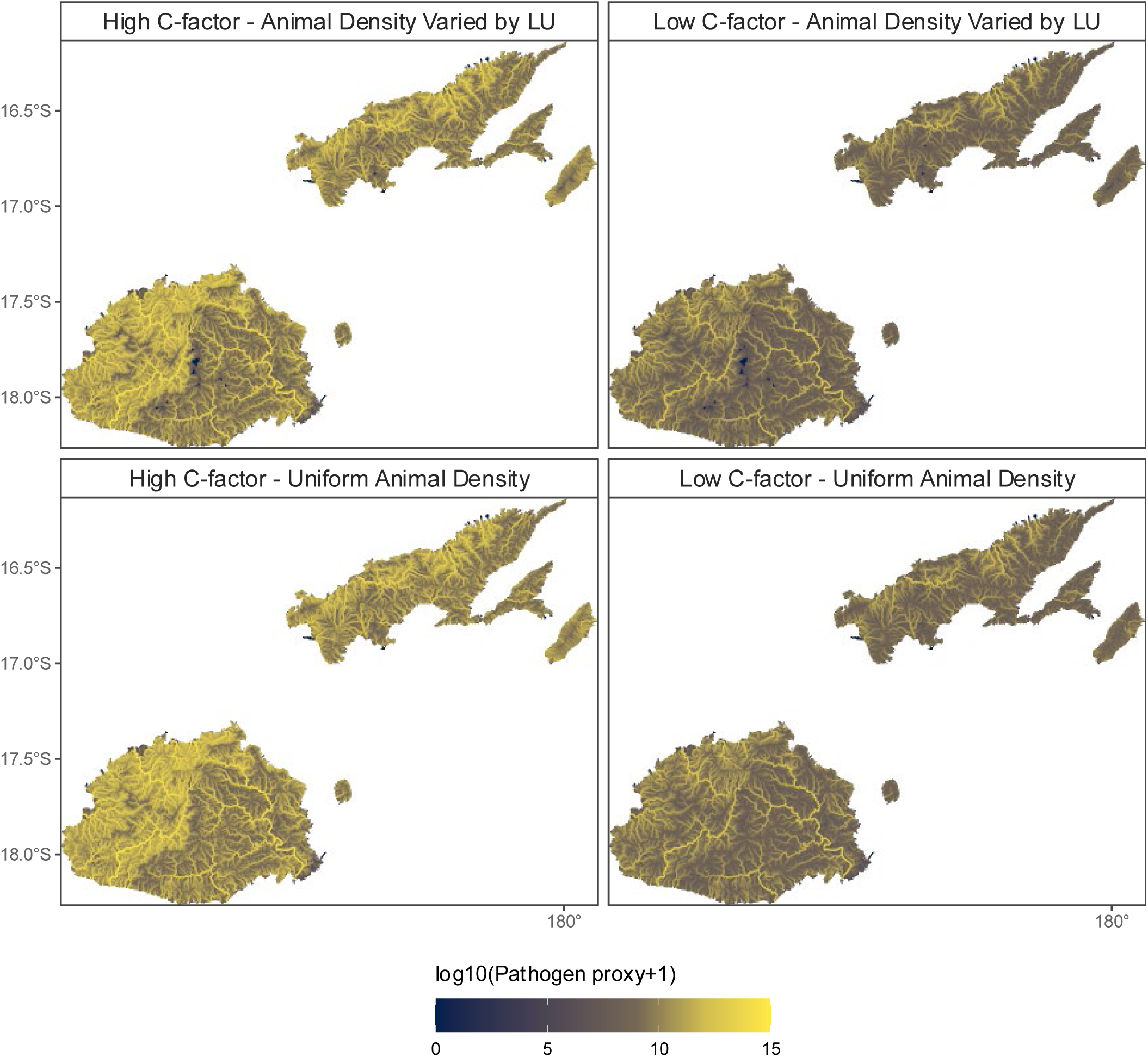
Log transformed accumulated pathogen load proxy calculated using four alternative assumptions: high C factor to calculate sediment export (left column), low C factor to calculate sediment export (right column), animal density varied using land use (i.e., varied; top row); and animal density not varied using land use (i.e., uniform; top row).

**Table A.** Results of univariate logistic regressions comparing four alternative accumulated pathogen loads with leptospirosis seropositivity. Proxy values were developed based on animal density estimates from varied or uniform land use, and sediment export estimates from high or low C factor. The value with the highest odds ratio (bolded) was included in the BN.

| Animal density | C factor | OR (95% confidence interval) | P value | AIC |
| --- | --- | --- | --- | --- |
| Varied by land use | High | 1.158 (1.063-1.261) | 0.0008 | 2119.6 |
|  | Low | 1.111 (1.007-1.226) | 0.0358 |  |
| Not varied by land use | High | 1.156 (1.063-1.257) | 0.0007 |  |
|  | Low | 1.112 (1.01-1.223) | 0.0293 |  |

**Table A.**
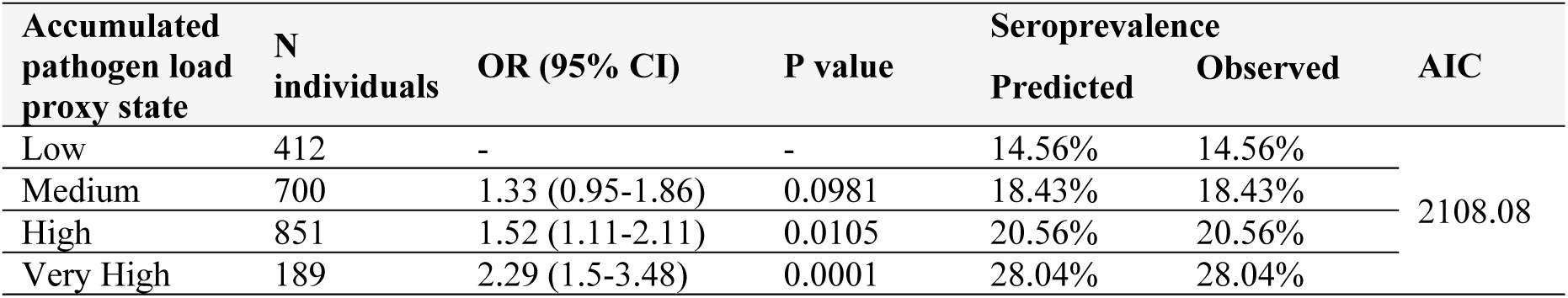
The result of a univariate logistic regression assessing the final accumulated pathogen load values used in the BN (developed based on animal density varied by land use, and sediment export estimates from high C factors), after discretisation into low, medium high, and very high states/categories.

**Fig C.**
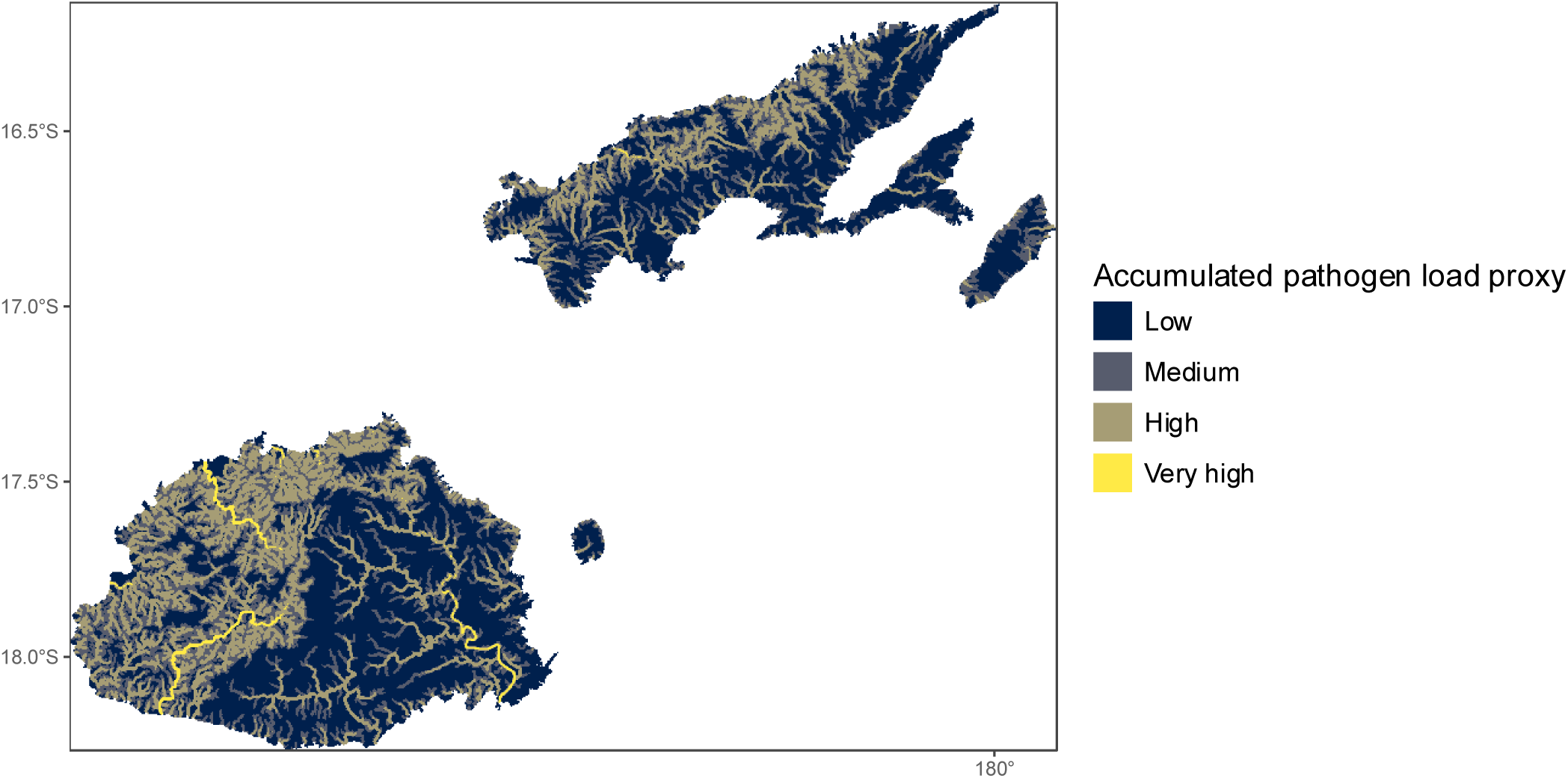
Discretised accumulated pathogen load proxy calculated using high C factor estimates and animal density varied using land use. Discretisation is into four states based on 50th, 75th, and 99th quantiles.

## S6 Appendix. Overview of Bayesian networks

BNs are a probabilistic modelling tool that uses Bayes’ theorem to assess and update the likelihood of an outcome given the influence of varying input variables (36). BNs are directed acyclic graphs with variables represented as nodes, joined by directional links (Fig A). Nodes exist in various states (e.g., working outdoors node having the states “Yes” and “No”; Fig A) and links point from a “parent node” to the “child node”, depicting conditional dependence. Conditional probability tables (CPTs) within each node quantify the chance of the node being in each possible state, which for child nodes, is conditional on the state of the parent node(s) (Fig A). The BN structure (i.e., nodes, links, states) and CPTs allows the calculation of influence through the network.

BNs are well suited to epidemiological modelling as they are able to incorporate a variety of data, assumptions, and sub-models (92). For example, CPTs can be populated using parameter learning algorithms from available data, and/or informed by expert opinion. Although links in a BN do not have to be causal, using causal relationships offers several benefits, including a direct representation of conditional independencies in the system, more intuitive conditional probabilities, and the ability to model the effects of interventions (93, 94).

**Fig A.**
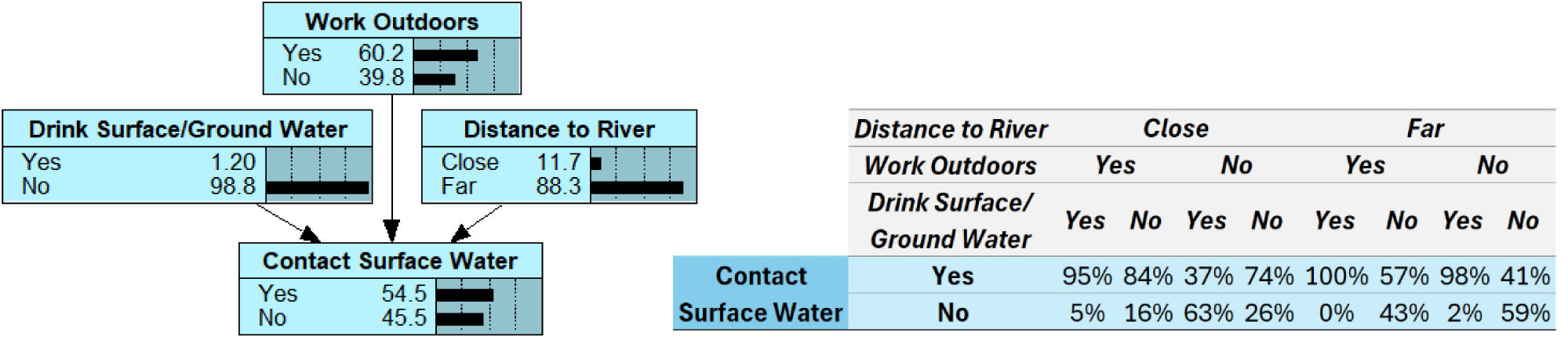
An example BN structure (left) and conditional probability table (right) estimating contact with surface water based on distance of household to river, outdoor work location, and drinking surface water or unprotected groundwater

## S7 Appendix. Parameterisation

We directly parameterised conditional probability tables (CPTs) where expert elicited data, national census results, or deterministic rules were available to provide a robust representation of conditions across Fiji. For example, the CPT for the *Exposure to Seropositive Cattle* node was parameterised with the logical relationship with *Cattle Seroprevalence* and *Exposure to Cattle* (Table E), and the CPT for the *Ethnicity* node was parameterised with ethnicity data available per tikina from the most recent population and household census. In cases where data were not known across all of Fiji, the probabilities were populated using EM. EM is a parameter learning algorithm used in BNs to populate incomplete CPTs or refine CPTs with values that maximize the likelihood of predicting observed data provided to the BN as a casefile (46). We created a casefile of 2,463 cases/individuals for the EM algorithm to learn from, which included socio-demographic, environmental, and serological data for individuals across two epidemiological surveys: 2,152 individuals in total from Lau et al. (15) and 311 individuals in total from Jupiter et al. (28).

### Initial parameterisation

Although the EM algorithm has been shown to provide robust predictions of CPTs when casefiles have sufficient data, it can generate spurious or extreme values for latent nodes which have no observed data to constrain predictions. As such, we initially parameterised latent nodes using coarse prior estimates which were then updated by the model during the learning phase (39). We guided the EM algorithm to refine latent node CPTs and prevented changes to fixed/set node CPTs by assigning “experience” within all CPTs (46).

Experience is the equivalent of a certain sample size observed for rows within the CPTs, i.e., confidence in the preset priors. Setting higher experience is equivalent to telling the model that the initial CPT parameterisation is derived from a large dataset, and therefore additional information presented within a casefile would cause little to no refinement of the CPT. Lower experience represents uncertain priors, and therefore EM learning may substantially change the CPT from initial values given information within a casefile. We excluded nodes with priors that were directly fixed/set (e.g., deterministic nodes) from EM learning, assigned some experience (i.e., 100) for latent nodes which were driven by priors that we wanted to be refined/updated, and no experience (i.e., 0) for priors with corresponding observed data to allow for robust predictions using our casefile.

To assist in developing initial probabilities in latent nodes with large CPTs and where unmodelled risk factors may be important (e.g., *Exposure to Water/Soil* node), we used a Noisy-OR model (39, 47). Within a BN, a Noisy-OR model^1^ uses the estimated independent influence of parent nodes (e.g., independent influence of *Working Outdoors* on *Exposure to Water/Soil* given all other drivers are not present) to interpolate a CPT using logic rules (95, 96). Importantly, the Noisy-OR model also includes a “leak” parameter that captures the influence of unmodelled causes, i.e., causes that are not explicitly included as parent nodes. We used the Noisy-OR model for seven latent nodes (e.g., Table A), allowing the consideration of unmodelled exposures to livestock (e.g., contact occurring outside of the household or community), unmodelled animal groups driving the direct infection, and unmodelled drivers of pathogenic *Leptospira* loads in the environment (e.g., if the person is exposed to many environments).

^1^ In the case of non-boolean nodes, the Noisy-OR model is generalised to Noisy-MAX (39, 47)

To assess the influence of initial latent node parameterisation on model discrimination and calibration, we developed three sets of different initial parameterisation values for latent node priors that the EM algorithm would update/revise:

- A “non-uniform” option where initial parameterisation of latent nodes differentiates the influence of parent nodes (e.g., *Contact with Surface Water* is more likely than *Outdoor Occupation* to cause *Exposure to Water / Soil*); and
- An “uniform” option where initial parameterisation of latent nodes does not differentiate between the influence of parent nodes (e.g., *Contact with Surface Water* is just as likely as *Outdoor Occupation* to cause *Exposure to Water / Soil*).
- A “control” initial parameterisation of latent nodes.

The initial parameterisation of latent nodes are provided below.

**Table A.** The Noisy-MAX values used to develop initial parameterisation of the CPT for the latent node representing exposure to water/soil. Green values represent initial parameterisation under non-uniform scenarios, and purple values represent the values used under uniform scenarios. In both cases, leak values were set assuming low influence of unmodelled risk factors, and some experience (i.e., 100) for all priors to guide expectation maximization algorithm in learning the final CPT, (*) except when drinking surface water confirmed, in which case the row was excluded from EM learning to reflect certainty of exposure under this situation and guide the algorithm away from changing this value. For control scenarios, no initial parameterisation values were used.

| Exposure to Water / Soil – non-uniform initial parameterisation |  |  |  |  |  |  |  |  |  |
| --- | --- | --- | --- | --- | --- | --- | --- | --- | --- |
|  | Contact with Flood Waters |  | Outdoor Work Location |  | Surface Water Contact |  | Drink Surface Water |  | LEAK |
|  | Yes | No | Yes | No | Yes | No | Yes* | No |  |
| Yes | 0.7 | 0 | 0.6 | 0 | 0.8 | 0 | 1 | 0 | 0.001 |
| No | 0.3 | 1 | 0.4 | 1 | 0.2 | 1 | 0 | 1 | 0.999 |
| Exposure to Water / Soil – uniform initial parameterisation |  |  |  |  |  |  |  |  |  |
|  | Contact with Flood Waters |  | Outdoor Work Location |  | Surface Water Contact |  | Drink Surface Water |  | LEAK |
|  | Yes | No | Yes | No | Yes | No | Yes* | No |  |
| Yes | 0.6 | 0 | 0.6 | 0 | 0.6 | 0 | 1 | 0 | 0.001 |
| No | 0.4 | 1 | 0.4 | 1 | 0.4 | 1 | 0 | 1 | 0.999 |

**Table B.**
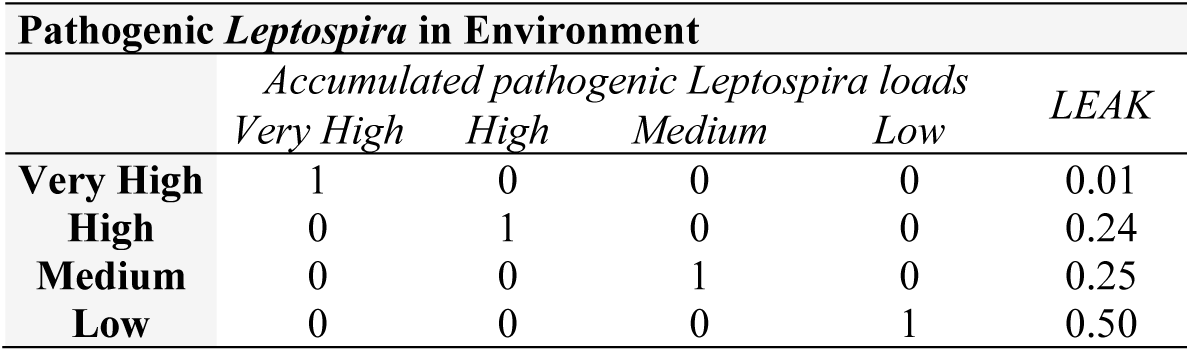
The Noisy-MAX values used to develop initial parameterisation of the CPT for the latent node representing pathogenic Leptospira in environment. Leak values were estimated using the percent coverage of the modelled accumulated pathogenic Leptospira load values across Fiji. Experience was set to 100 for all priors to guide expectation maximization algorithm in learning the final CPT.

**Table C.** The Noisy-MAX values used to develop initial parameterisation of the CPTs for the latent nodes representing contact with cattle, pigs, horses, and goats. Green values represent initial parameterisation under non-uniform scenarios, and purple values represent the values used under uniform scenarios. In both cases, leak values were set assuming low influence of unmodelled risk factors, and experience was set to 100 for all priors to guide expectation maximization algorithm in learning the final CPTs. For control scenarios, no initial parameterisation values were used.

| Contact with [Cattle / Pigs / Horses / Goats] – non-uniform initial parameterisation |  |  |  |  |  |
| --- | --- | --- | --- | --- | --- |
|  | [Cattle / Pigs / Horses / Goats] in Household |  | [Cattle / Pigs / Horses / Goats] in Community |  | LEAK |
|  | Yes | No | Yes | No |  |
| Yes | 0.8 | 0 | 0.6 | 0 | 0.001 |
| No | 0.2 | 1 | 0.4 | 1 | 0.999 |
| Contact with [Cattle / Pigs / Horses / Goats] – uniform initial parameterisation |  |  |  |  |  |
|  | [Cattle / Pigs / Horses / Goats] in Household |  | [Cattle / Pigs / Horses / Goats] in Community |  | LEAK |
|  | Yes | No | Yes | No |  |
| Yes | 0.6 | 0 | 0.6 | 0 | 0.001 |
| No | 0.4 | 1 | 0.4 | 1 | 0.999 |

**Table D.** CPT for nodes representing leptospirosis seroprevalence in cattle, pigs, horses, goats, and rodents (five separate nodes). Probabilities were extracted from a triangular distribution developed from expert-elicited best, lowest, and highest estimates. These nodes were excluded from EM learning.

| Seroprevalence of Leptospirosis |  |  |  |  |  |
| --- | --- | --- | --- | --- | --- |
|  | In Cattle | In Pigs | In Horses | In Goats | In Rodents |
| 0-10% | 0 | 0.04 | 0 | 0.08 | 0 |
| 10-20% | 0.01 | 0.29 | 0 | 0.41 | 0 |
| 20-30% | 0.12 | 0.36 | 0 | 0.39 | 0.13 |
| 30-40% | 0.28 | 0.22 | 0.05 | 0.12 | 0.37 |
| 40-50% | 0.38 | 0.09 | 0.19 | 0 | 0.37 |
| 50-60% | 0.20 | 0 | 0.33 | 0 | 0.13 |
| 60-70% | 0.02 | 0 | 0.33 | 0 | 0 |
| 70-80% | 0 | 0 | 0.11 | 0 | 0 |
| 80-90% | 0 | 0 | 0 | 0 | 0 |
| 90-100% | 0 | 0 | 0 | 0 | 0 |

**Table E.** CPT for nodes representing exposure to infected cattle, pigs, horses, goats, and rodents (five separate nodes). Parameterisation is identical across all five nodes and is determined using seroprevalence of leptospirosis in the animal population when exposure to the animal is present, and otherwise the probability is zero as there is no exposure. These nodes were excluded from EM learning.

| Exposure to Infected [Cattle / Pigs / Horses / Goats / Rodents] |  |  |  |  |  |  |  |  |  |  |  |  |  |  |  |  |  |  |  |  |
| --- | --- | --- | --- | --- | --- | --- | --- | --- | --- | --- | --- | --- | --- | --- | --- | --- | --- | --- | --- | --- |
|  | Yes |  |  |  |  |  |  |  |  |  | No |  |  |  |  |  |  |  |  |  |
|  | Exposure to [Cattle / Pigs / Horses / Goats / Rodents] |  |  |  |  |  |  |  |  |  | Exposure to [Cattle / Pigs / Horses / Goats / Rodents] |  |  |  |  |  |  |  |  |  |
|  | Seroprevalence in [Cattle / Pigs / Horses / Goats / Rodents] (%) |  |  |  |  |  |  |  |  |  | Seroprevalence in [Cattle / Pigs / Horses / Goats / Rodents] (%) |  |  |  |  |  |  |  |  |  |
|  | 0-10 | 10-20 | 20-30 | 30-40 | 40-50 | 50-60 | 60-70 | 70-80 | 80-90 | 90-100 | 0-10 | 10-20 | 20-30 | 30-40 | 40-50 | 50-60 | 60-70 | 70-80 | 80-90 | 90-100 |
| Yes | 0.1 | 0.2 | 0.3 | 0.4 | 0.5 | 0.6 | 0.7 | 0.8 | 0.9 | 0.99 | 0 | 0 | 0 | 0 | 0 | 0 | 0 | 0 | 0 | 0 |
| No | 0.9 | 0.8 | 0.7 | 0.6 | 0.5 | 0.4 | 0.3 | 0.2 | 0.1 | 0.01 | 1 | 1 | 1 | 1 | 1 | 1 | 1 | 1 | 1 | 1 |

**Table F.**
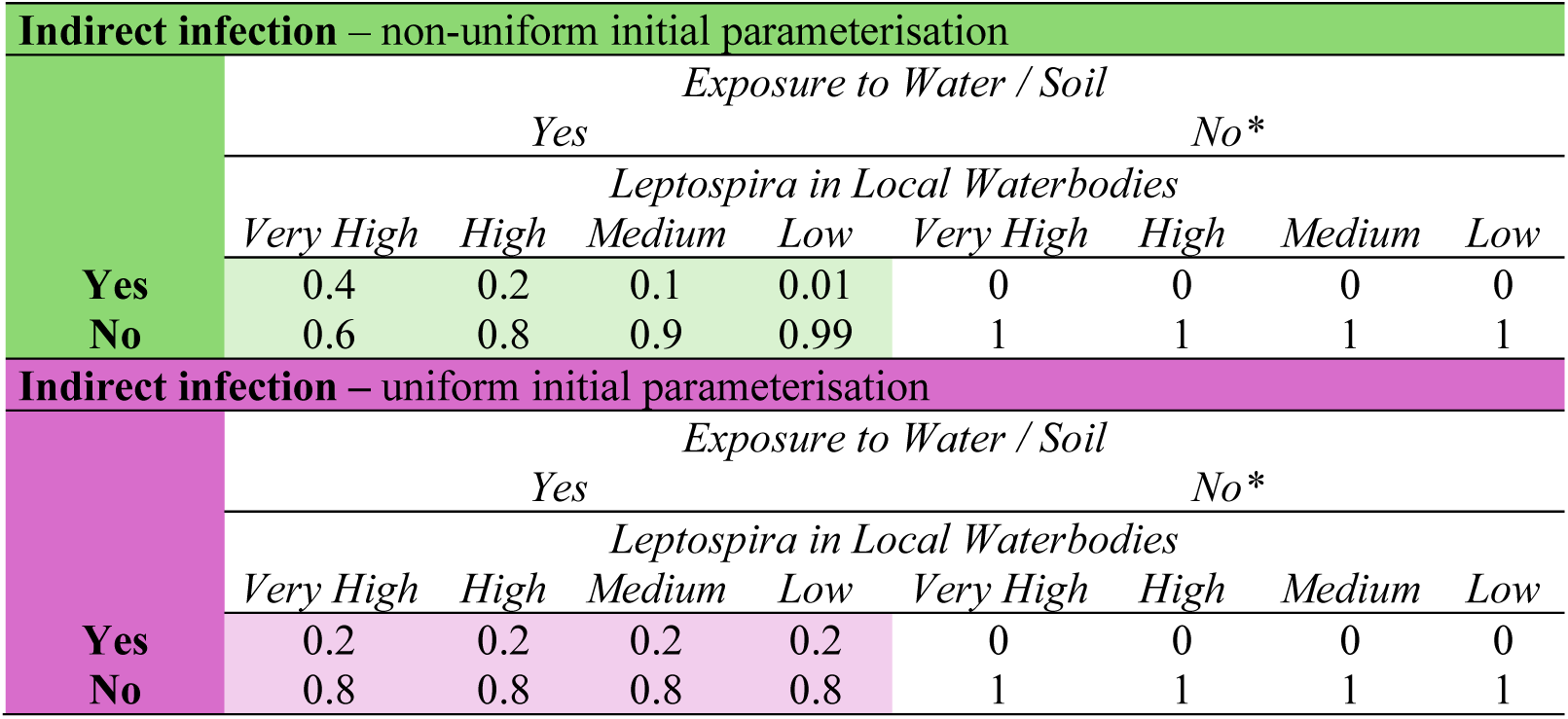
The initial parameterisation of the CPT for the latent node representing indirect infection from the environment. Green values represent initial parameterisation under non-uniform scenarios, and purple values represent the values used under uniform scenarios. In both cases, experience was set to 100 for all priors to guide expectation maximization algorithm in learning the final CPT, (*) except when there was no exposure to water/soil, in which case the row was excluded from EM learning to reflect certainty of no infection under this situation and guide the algorithm away from changing this value. For control scenarios, no initial parameterisation values were used.

**Table G.** The Noisy-MAX values used to develop initial parameterisation of the CPTs for the latent nodes representing direct infection from animals. Green values represent initial parameterisation under non-uniform scenarios, and purple values represent the values used under uniform scenarios. In both cases, leak values were set assuming low influence of unmodelled risk factors, and experience was set to 100 for all priors to guide expectation maximization algorithm in learning the final CPTs. For control scenarios, no initial parameterisation values were used.

| Direct Infection– non-uniform initial parameterisation |  |  |  |  |  |  |  |  |  |  |  |
| --- | --- | --- | --- | --- | --- | --- | --- | --- | --- | --- | --- |
|  | Exposure to infected Cattle |  | Exposure to infected Pigs |  | Exposure to infected Goats |  | Exposure to infected Horses |  | Exposure to infected Rodents |  | LEAK |
|  | Yes | No | Yes | No | Yes | No | Yes | No | Yes | No |  |
| Yes | 0.15 | 0 | 0.15 | 0 | 0.15 | 0 | 0.15 | 0 | 0.2 | 0 | 0.001 |
| No | 0.85 | 1 | 0.85 | 1 | 0.85 | 1 | 0.85 | 1 | 0.8 | 1 | 0.999 |
| Direct Infection– uniform initial parameterisation |  |  |  |  |  |  |  |  |  |  |  |
|  | Exposure to infected Cattle |  | Exposure to infected Pigs |  | Exposure to infected Goats |  | Exposure to infected Horses |  | Exposure to infected Rodents |  | LEAK |
|  | Yes | No | Yes | No | Yes | No | Yes | No | Yes | No |  |
| Yes | 0.2 | 0 | 0.2 | 0 | 0.2 | 0 | 0.2 | 0 | 0.2 | 0 | 0.001 |
| No | 0.8 | 1 | 0.8 | 1 | 0.8 | 1 | 0.8 | 1 | 0.8 | 1 | 0.999 |

---

**Fig S1.**
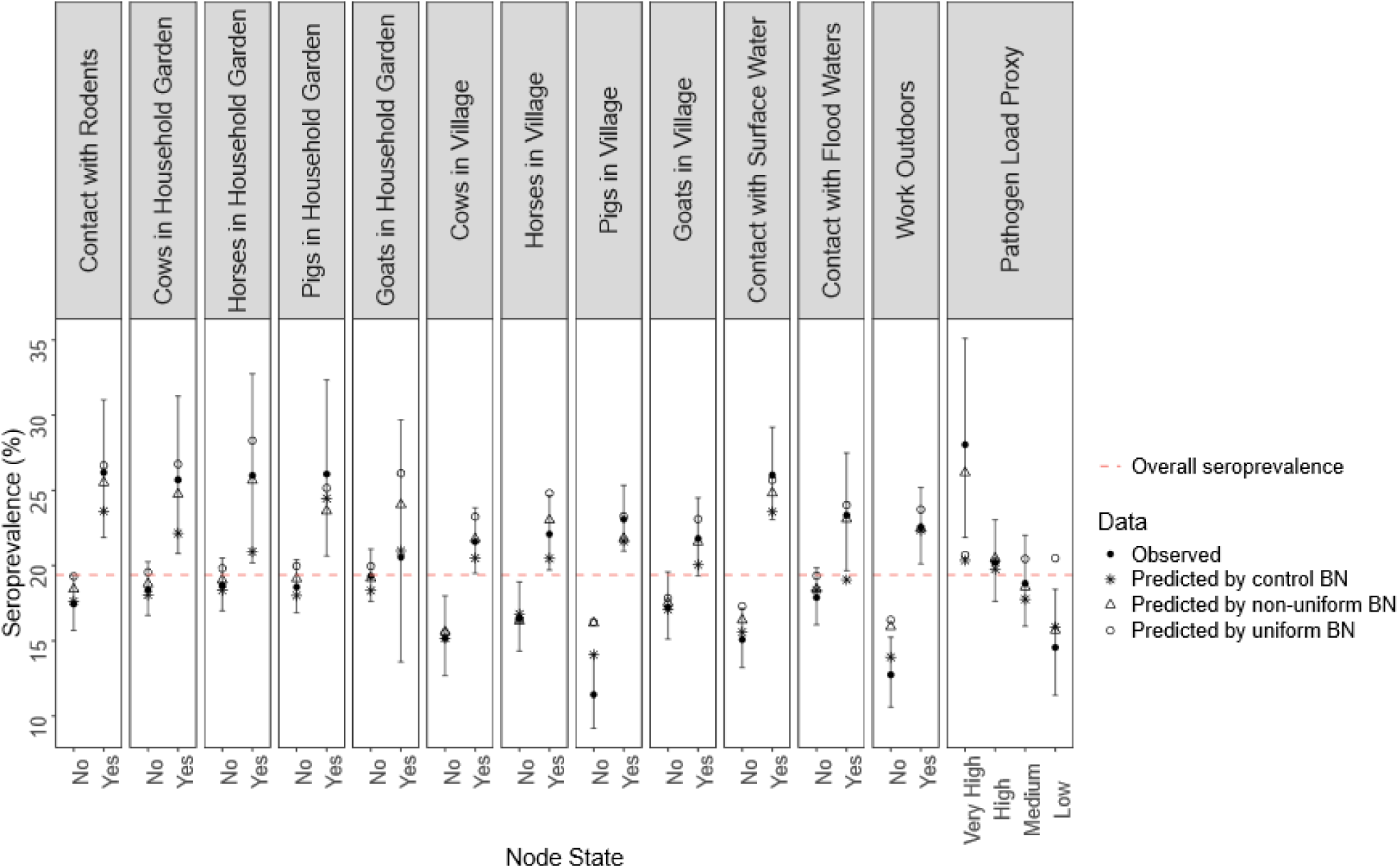
Comparison between observed and Bayesian Network (BN) predicted leptospirosis seroprevalence for individuals exposed to risk factor scenarios of interest. Black points are observed values (plotted with 95% confidence interval), asterisks are control BN predictions, triangles are non-uniform BN predictions, and circles are uniform BN predictions. The red dotted line displays the observed seroprevalence across all individuals.

**Table S1.**
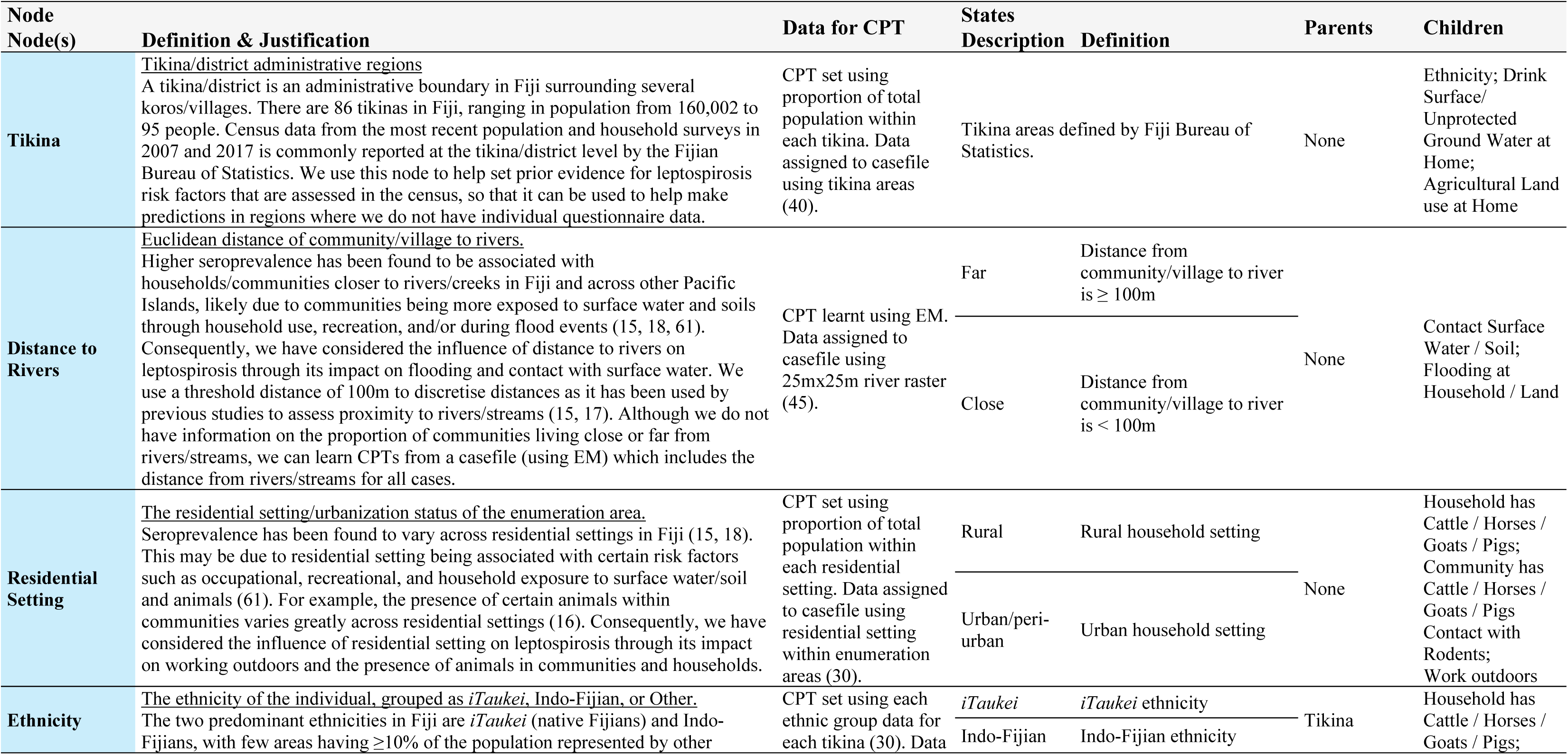
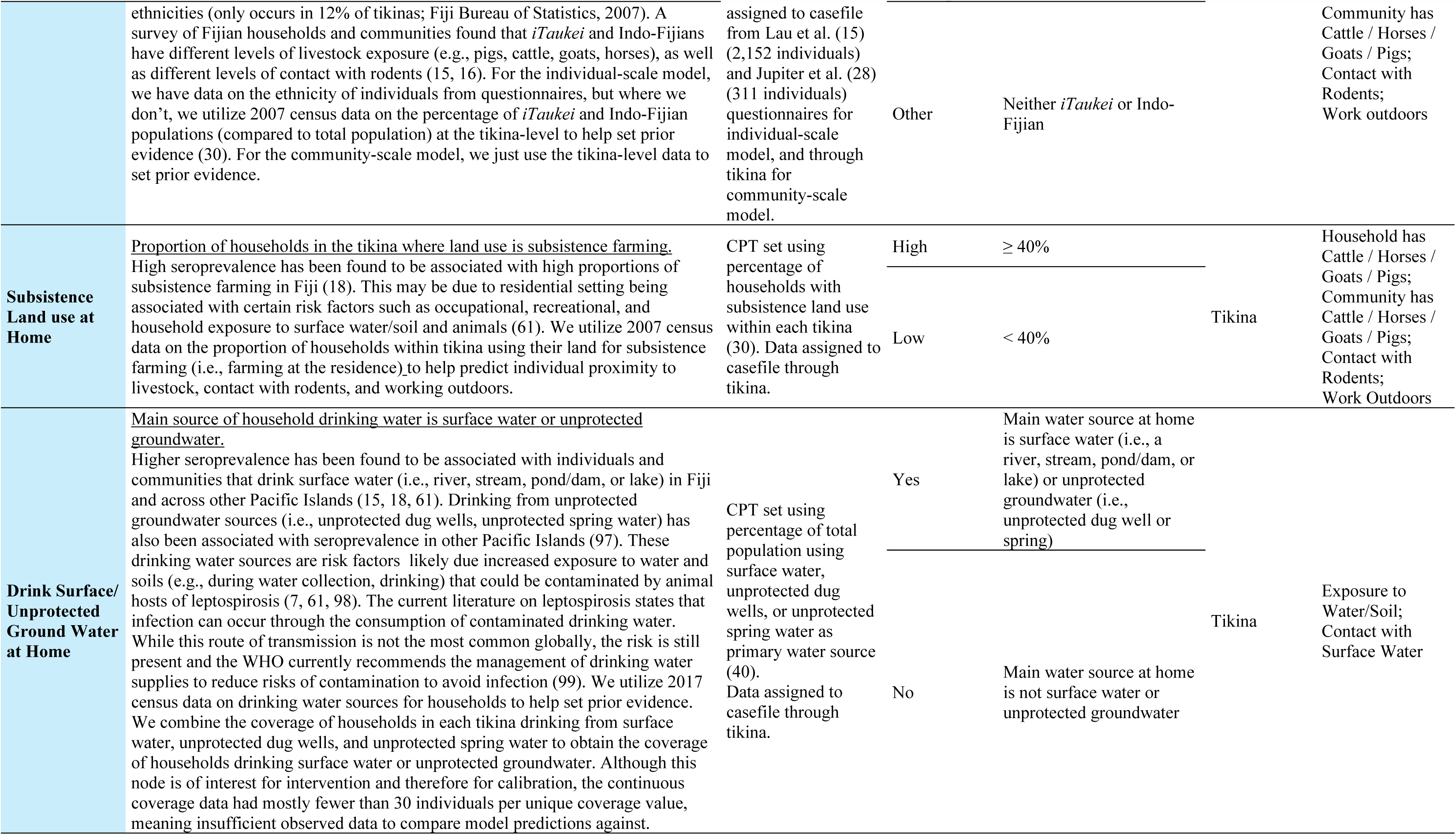
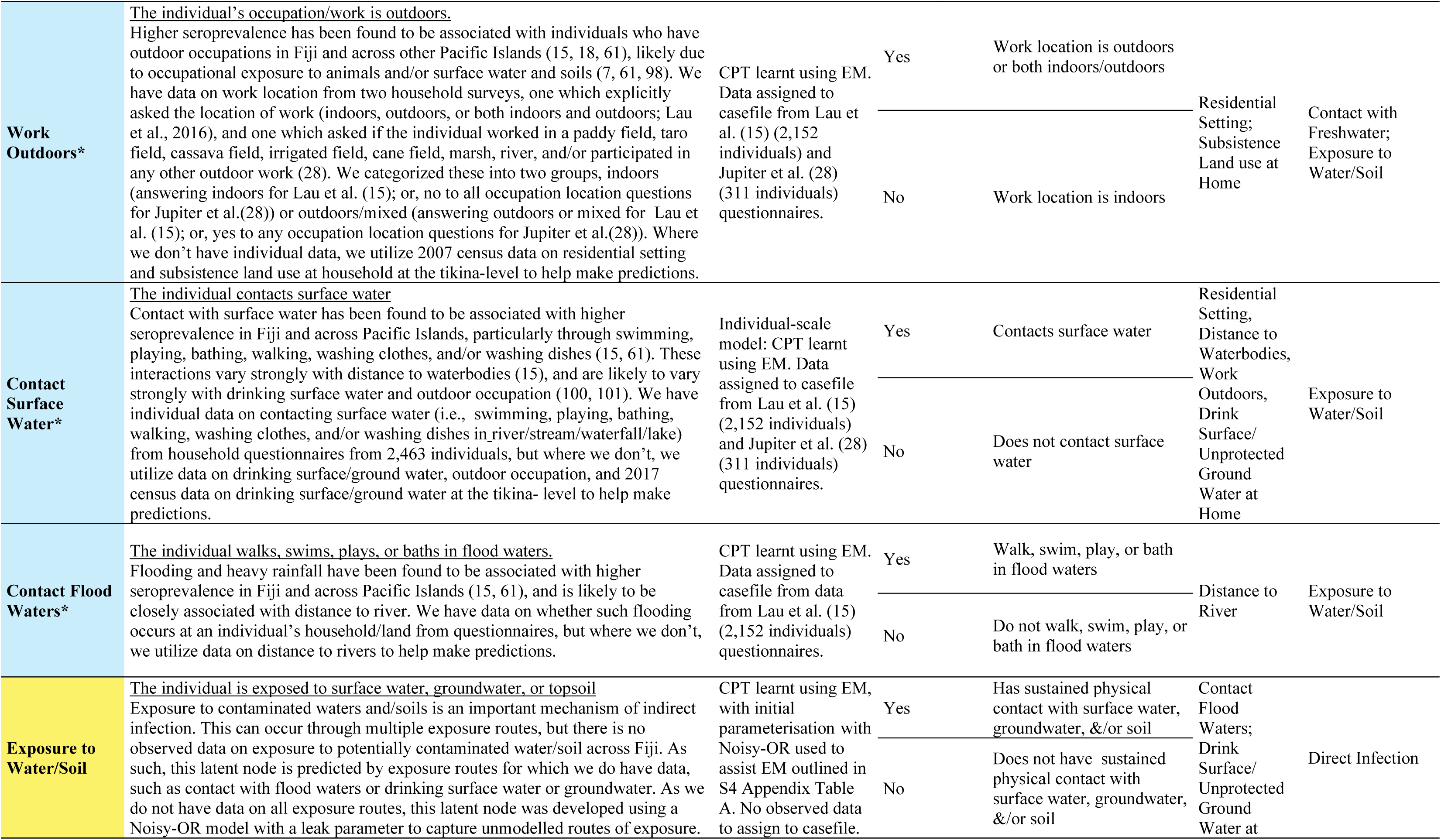
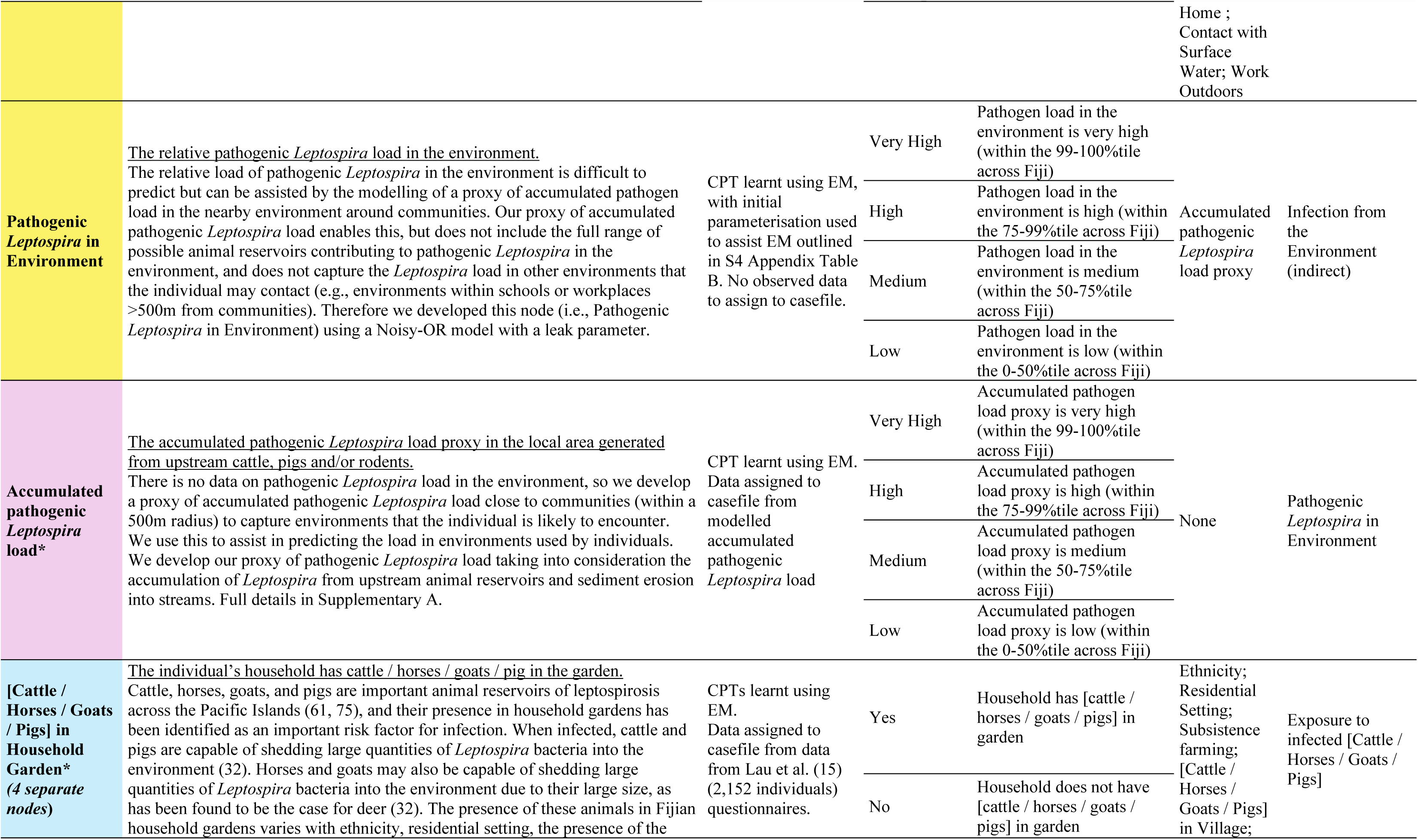
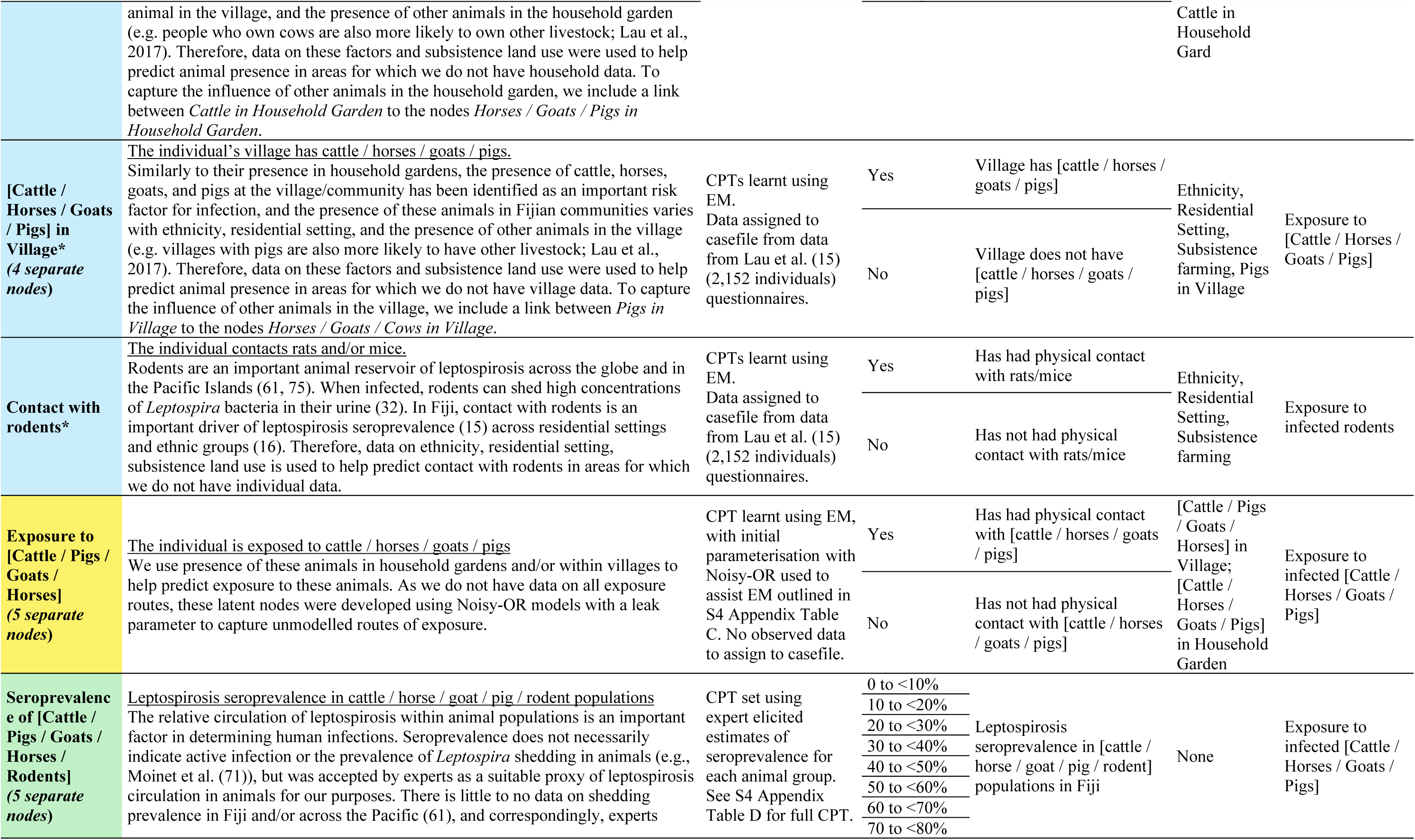
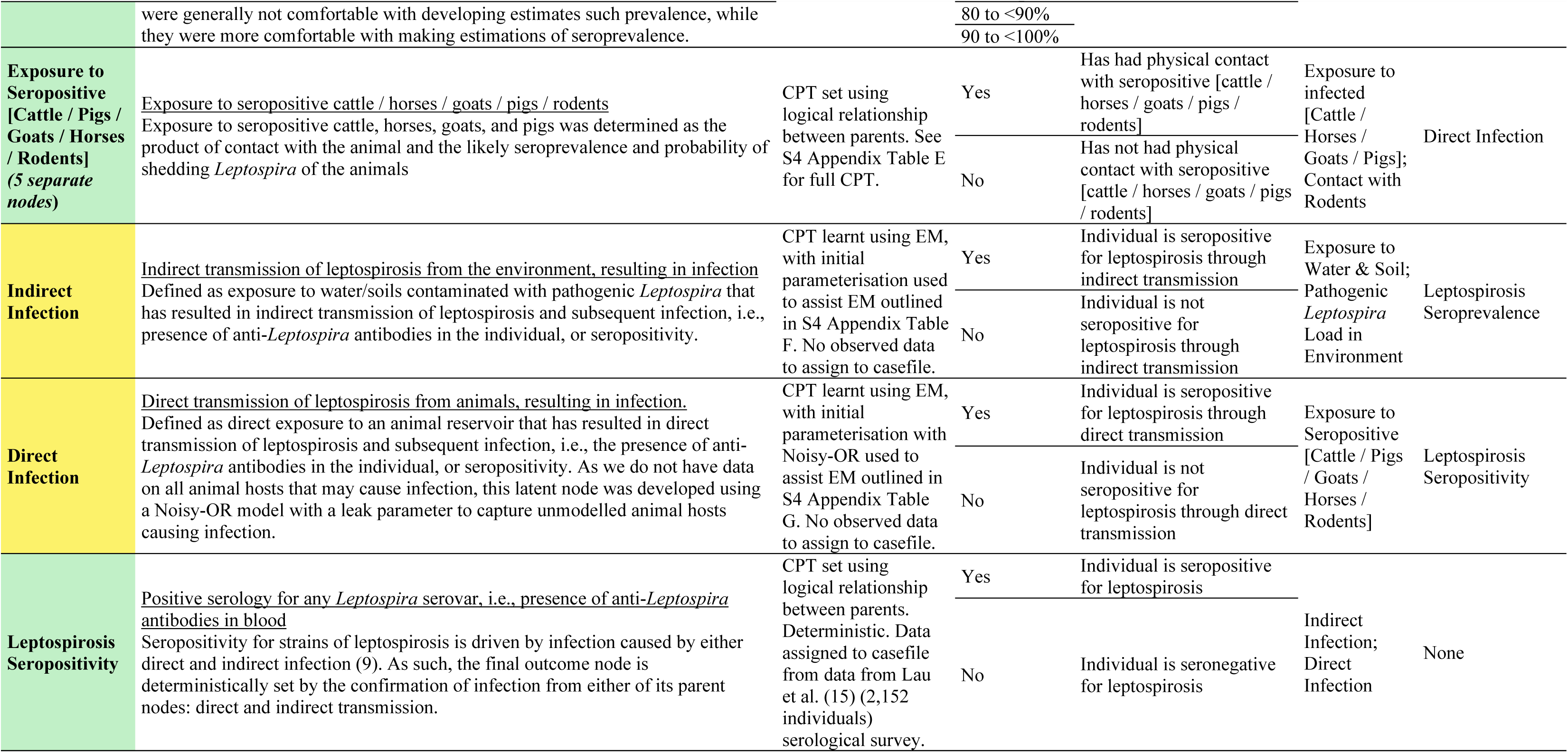
The nodes, states, data sources for learning conditional probability tables (CPT), and links/arcs included in the leptospirosis infection Bayesian network for Fiji. Blue represents nodes parameterised by data, purple represents the node parametrised by the hydrologically linked pathogen model, green represents nodes parameterised by expert elicitation, and yellow represents latent nodes learnt via EM with guidance from initial parameterisation. * indicates the node was selected for model calibration validity testing.

**Table S2.**
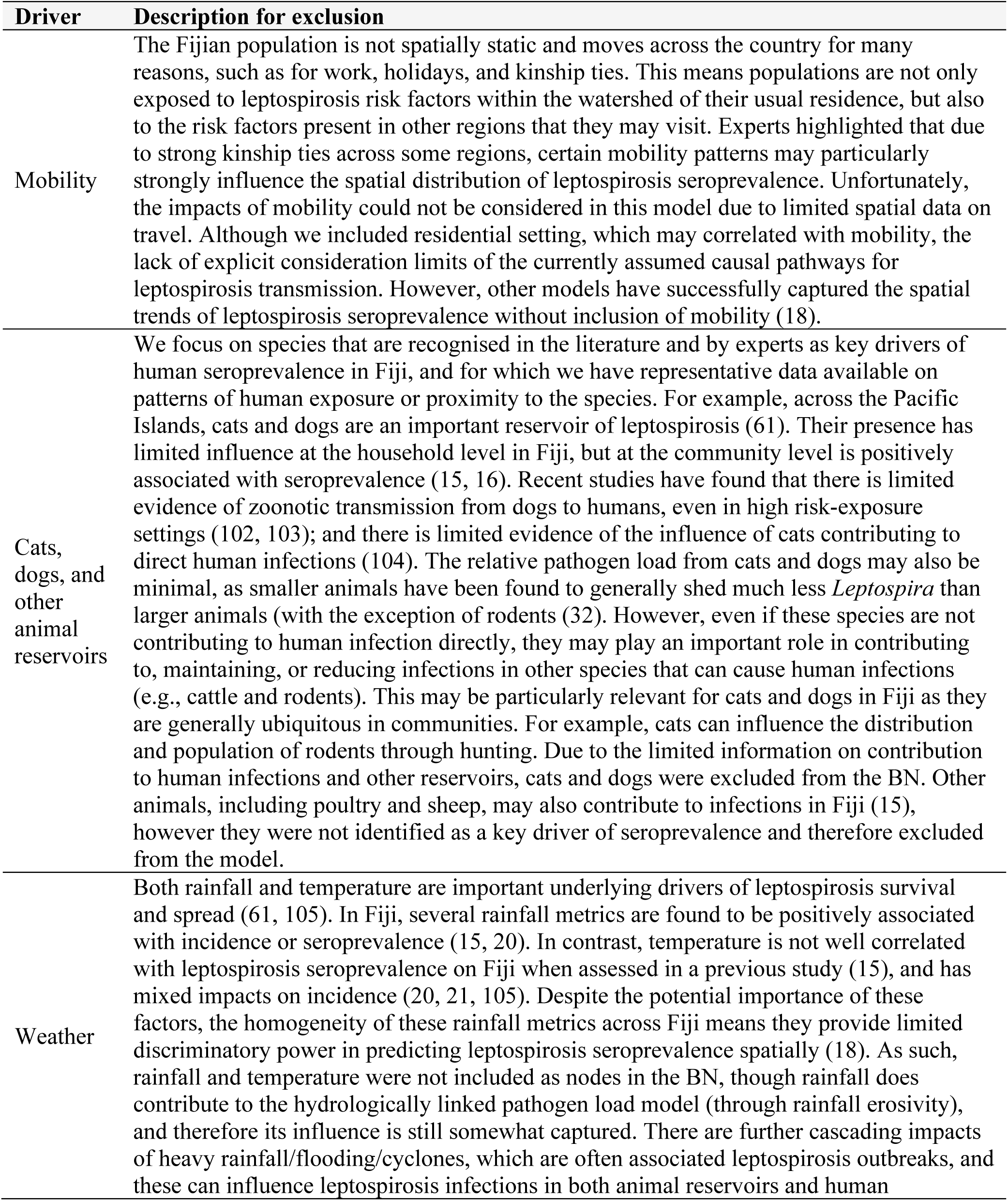
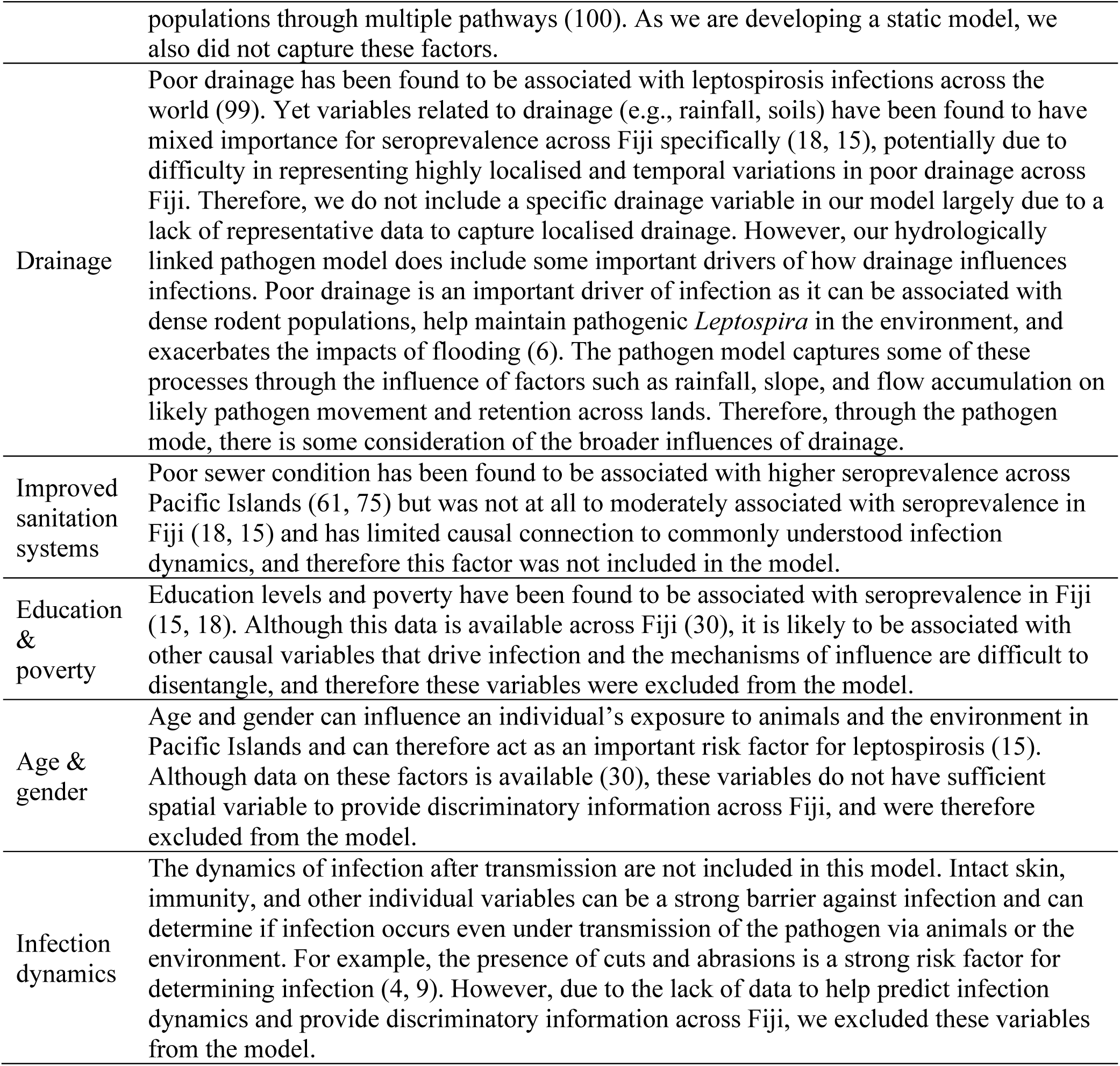
Potentially important leptospirosis risk factors identified by experts or within literature yet excluded from model and reason for exclusion

**Table S3.**
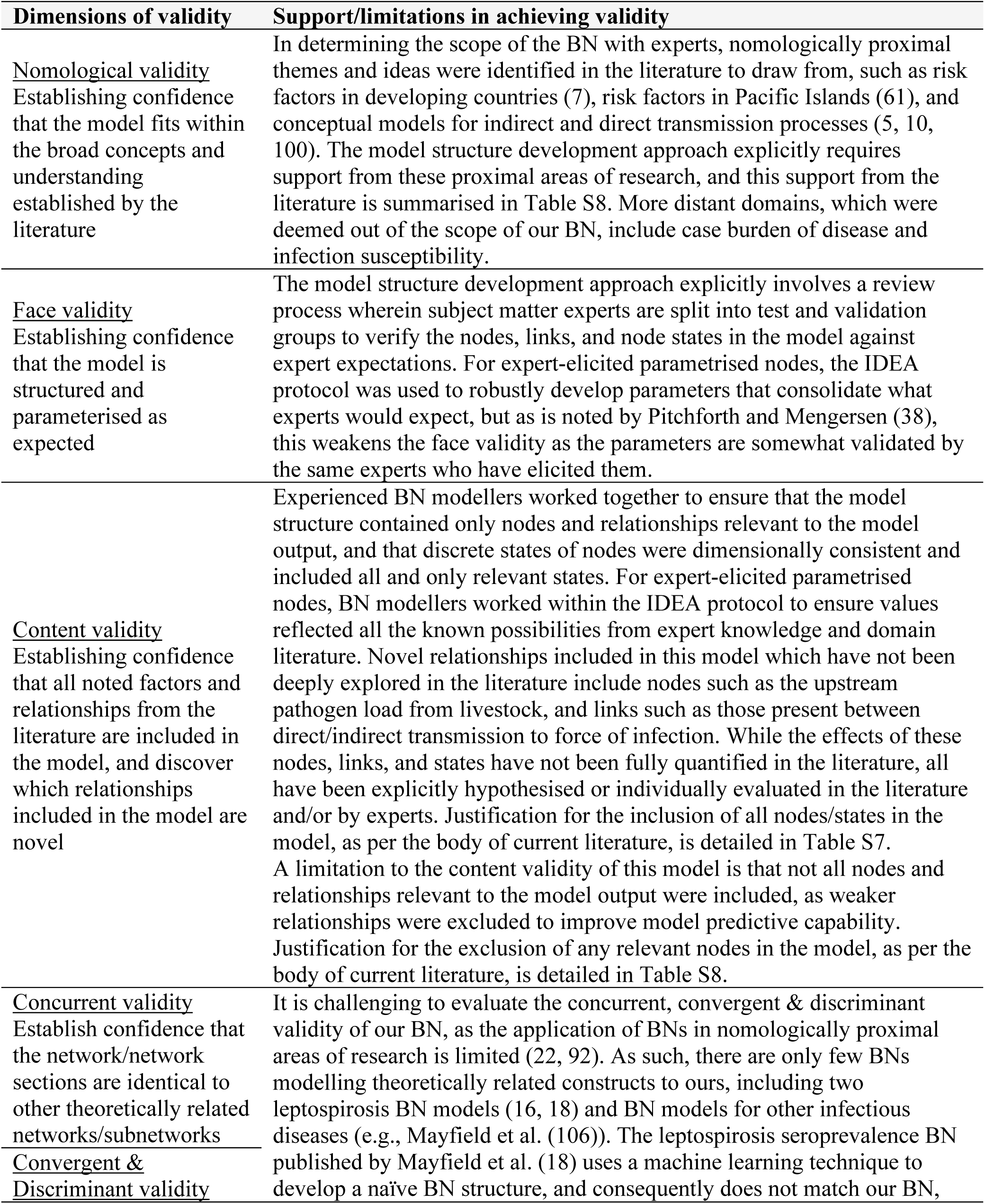
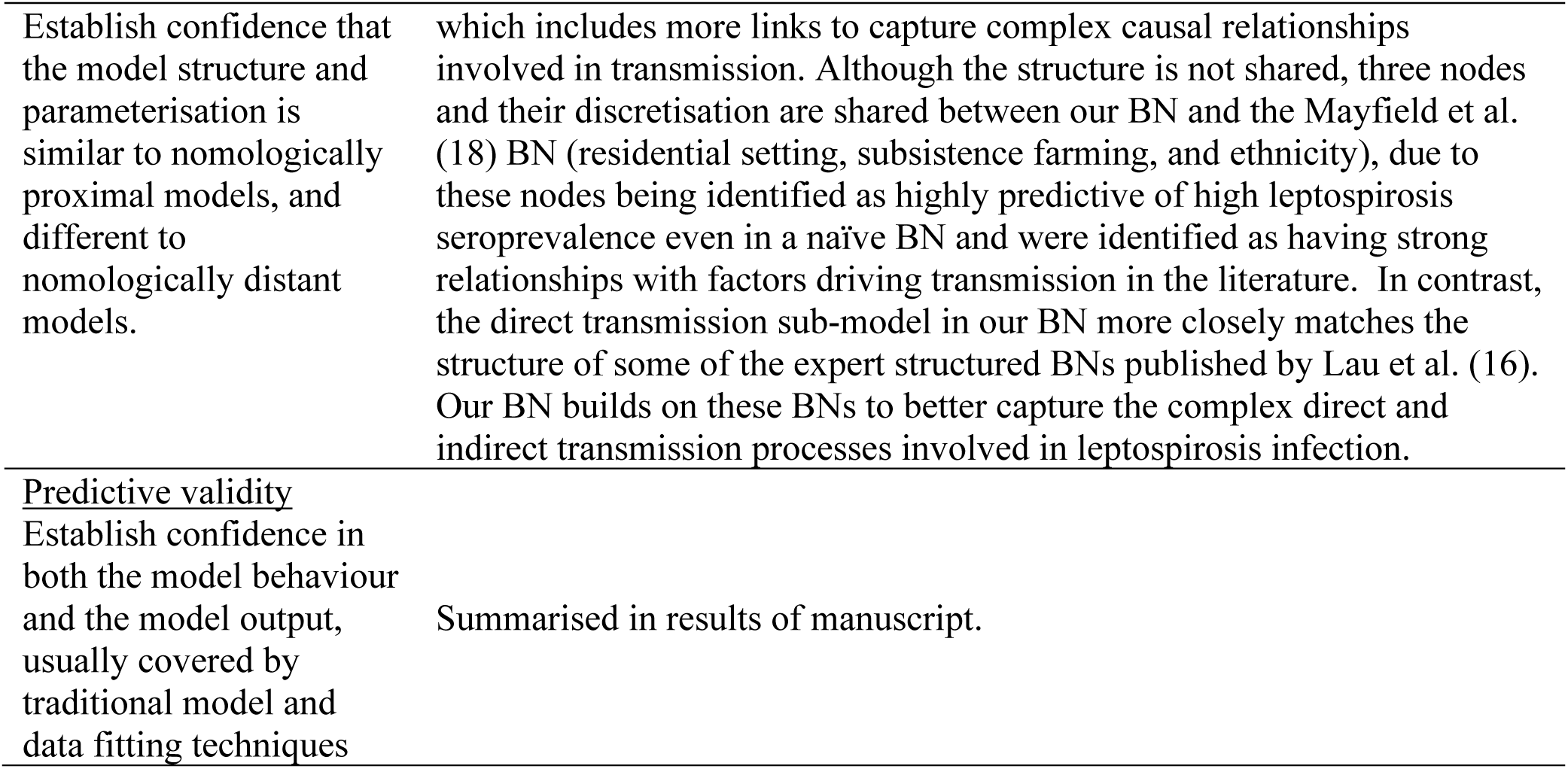
Validity assessment of the Bayesian Network, based on the validity testing framework described in Pitchforth and Mengersen (38).

**Table S4.** Comparison of discrimination capability (measured as AUC) and the predicted probability of direct and indirect leptospirosis infections between three different hybrid models of leptospirosis.

| Latent node<br>initial<br>parameterisation | Model AUC<sup>^</sup><br>median (IQR) | Predicted probability of infection* |  |  |
| --- | --- | --- | --- | --- |
|  |  | Indirect infection | Direct infection | Leptospirosis<br>Seropositivity |
| **Non-uniform** | 0.62 (0.62 - 0.63) | 9.09% | 11.5% | 19.5% |
| **Uniform** | 0.62 (0.61 - 0.63) | 8.96% | 12.6% | 20.4% |
| **Control** | 0.62 (0.62-0.63) | 7.93% | 11.5% | 18.5% |

<sup>^</sup> AUC values summarise model discrimination over ten trials, each using a randomly assigned 50% of the casefile data for training and the remaining 50% for validating
\* Predicted probabilities of infection are obtained from hybrid models trained on the full casefile data
IQR = interquartile range

